## Supplemental Tables 1-12 for "Hospital segregation, critical care strain, and inpatient mortality during the COVID-19 pandemic in New York City"

Table S1: Patient and admission characteristics of overall hospitalizations among adults (18+) in NYC by hospital quartile, January 5-May 9, 2020[a]

| Characteristic, No. (%) | NYC (N=47 hospitals)[b] | | Quartile 1 (lowest % of Medicaid or Self-pay patients) (N=11 hospitals) | | Quartile 2 (N=12 hospitals) | | Quartile 3 (N=12 hospitals) | | Quartile 4 (highest % of Medicaid or Self-pay patients) (N=12 hospitals) | |
|---|---|---|---|---|---|---|---|---|---|---|
| | **Pre-pandemic[c]** | **Wave 1[d]** | **Pre-pandemic** | **Wave 1** | **Pre-pandemic** | **Wave 1** | **Pre-pandemic** | **Wave 1** | **Pre-pandemic** | **Wave 1** |
| Overall hospitalizations | 145,054 (100%) | 120,275 (100%) | 49,977 (100%) | 36,257 (100%) | 38,360 (100%) | 32,062 (100%) | 29,846 (100%) | 26,020 (100%) | 26,871 (100%) | 25,936 (100%) |
| Elixhauser comorbidity score, mean (SD) | 6.3 (9.3) | 6.7 (8.8) | 6.8 (9.4) | 7.1 (9.1) | 7.1 (9.3) | 7.3 (8.8) | 5.9 (9.2) | 6.6 (8.6) | 4.8 (8.9) | 5.5 (8.5) |
| Nursing home admission | 6,204 (4.3%) | 8,037 (6.7%) | 1,232 (2.5%) | 1,345 (3.7%) | 2,449 (6.4%) | 3,066 (9.6%) | 2,009 (6.7%) | 2,804 (11%) | 514 (1.9%) | 822 (3.2%) |
| Emergency department | 100,604 (69%) | 95,953 (80%) | 30,064 (60%) | 26,682 (74%) | 25,893 (68%) | 26,033 (81%) | 23,395 (78%) | 22,210 (85%) | 21,252 (79%) | 21,028 (81%) |
| Major SOI present on admission[e] | 30,368 (21%) | 25,057 (21%) | 10,841 (22%) | 7,999 (22%) | 8,617 (22%) | 7,022 (22%) | 5,862 (20%) | 5,457 (21%) | 5,048 (19%) | 4,579 (18%) |
| Extreme SOI present on admission | 9,412 (6.5%) | 11,954 (9.9%) | 3,228 (6.5%) | 3,396 (9.4%) | 2,749 (7.2%) | 3,384 (11%) | 2,032 (6.8%) | 2,805 (11%) | 1,403 (5.2%) | 2,369 (9.1%) |
| Gender | | | | | | | | | | |
| Female | 80,784 (56%) | 61,672 (51%) | 28,821 (58%) | 20,249 (56%) | 22,008 (57%) | 16,633 (52%) | 16,417 (55%) | 13,051 (50%) | 13,538 (50%) | 11,739 (45%) |
| Male | 64,255 (44%) | 58,591 (49%) | 21,153 (42%) | 16,001 (44%) | 16,352 (43%) | 15,429 (48%) | 13,428 (45%) | 12,969 (50%) | 13,322 (50%) | 14,192 (55%) |
| Age group | | | | | | | | | | |
| Age 18-39 | 38,674 (27%) | 29,762 (25%) | 13,159 (26%) | 9,932 (27%) | 8,550 (22%) | 6,852 (21%) | 8,389 (28%) | 6,298 (24%) | 8,576 (32%) | 6,680 (26%) |
| Age 40-64 | 50,162 (35%) | 41,962 (35%) | 15,803 (32%) | 11,405 (31%) | 12,891 (34%) | 11,225 (35%) | 10,174 (34%) | 8,653 (33%) | 11,294 (42%) | 10,679 (41%) |
| Age 65-79 | 34,472 (24%) | 30,698 (26%) | 12,940 (26%) | 9,317 (26%) | 10,181 (27%) | 8,780 (27%) | 6,481 (22%) | 6,653 (26%) | 4,870 (18%) | 5,948 (23%) |
| Age 80+ | 21,746 (15%) | 17,853 (15%) | 8,075 (16%) | 5,603 (15%) | 6,738 (18%) | 5,205 (16%) | 4,802 (16%) | 4,416 (17%) | 2,131 (7.9%) | 2,629 (10%) |
| Race/ethnicity | | | | | | | | | | |
| Hispanic | 35,031 (24%) | 32,027 (27%) | 8,274 (17%) | 6,917 (19%) | 9,585 (25%) | 8,835 (28%) | 7,198 (24%) | 6,711 (26%) | 9,974 (37%) | 9,564 (37%) |
| NH Asian or Pacific Islander | 10,313 (7.1%) | 8,788 (7.3%) | 3,043 (6.1%) | 2,425 (6.7%) | 3,481 (9.1%) | 2,862 (8.9%) | 2,669 (8.9%) | 2,414 (9.3%) | 1,120 (4.2%) | 1,087 (4.2%) |
| NH Black | 39,529 (27%) | 33,021 (27%) | 9,478 (19%) | 7,204 (20%) | 11,472 (30%) | 9,569 (30%) | 8,040 (27%) | 6,573 (25%) | 10,539 (39%) | 9,675 (37%) |
| NH White | 42,027 (29%) | 29,938 (25%) | 22,541 (45%) | 14,563 (40%) | 9,337 (24%) | 6,728 (21%) | 8,316 (28%) | 6,967 (27%) | 1,833 (6.8%) | 1,680 (6.5%) |
| Other | 18,154 (13%) | 16,501 (14%) | 6,641 (13%) | 5,148 (14%) | 4,485 (12%) | 4,068 (13%) | 3,623 (12%) | 3,355 (13%) | 3,405 (13%) | 3,930 (15%) |
| Insurance | | | | | | | | | | |
| Medicaid | 50,302 (35%) | 41,192 (34%) | 10,380 (21%) | 8,247 (23%) | 11,494 (30%) | 9,807 (31%) | 13,191 (44%) | 10,556 (41%) | 15,237 (57%) | 12,582 (49%) |
| Medicare | 61,027 (42%) | 51,073 (42%) | 22,682 (45%) | 15,769 (43%) | 18,557 (48%) | 14,731 (46%) | 12,213 (41%) | 11,572 (44%) | 7,575 (28%) | 9,001 (35%) |
| Private insurance | 29,596 (20%) | 25,217 (21%) | 15,414 (31%) | 11,365 (31%) | 7,768 (20%) | 7,063 (22%) | 3,693 (12%) | 3,333 (13%) | 2,721 (10%) | 3,456 (13%) |
| Self-pay | 2,757 (1.9%) | 2,050 (1.7%) | 754 (1.5%) | 536 (1.5%) | 303 (0.8%) | 240 (0.7%) | 501 (1.7%) | 450 (1.7%) | 1,199 (4.5%) | 824 (3.2%) |
| Other | 1,372 (0.9%) | 743 (0.6%) | 747 (1.5%) | 340 (0.9%) | 238 (0.6%) | 221 (0.7%) | 248 (0.8%) | 109 (0.4%) | 139 (0.5%) | 73 (0.3%) |
| Neighborhood poverty | | | | | | | | | | |
| 0 to <10% | 23,104 (18%) | 17,968 (16%) | 11,383 (27%) | 8,080 (25%) | 7,419 (21%) | 6,056 (20%) | 2,346 (8.1%) | 1,962 (7.7%) | 1,956 (7.5%) | 1,870 (7.3%) |

| | | | | | | | | | | |
|---|---|---|---|---|---|---|---|---|---|---|
| 10 to <20% | 54,704 (42%) | 50,491 (44%) | 16,789 (40%) | 13,417 (41%) | 15,551 (45%) | 14,914 (50%) | 13,453 (46%) | 12,326 (48%) | 8,911 (34%) | 9,834 (39%) |
| 20 to <30% | 29,730 (23%) | 25,332 (22%) | 8,977 (21%) | 7,300 (22%) | 6,461 (19%) | 5,316 (18%) | 8,483 (29%) | 7,591 (30%) | 5,809 (22%) | 5,125 (20%) |
| 30% or higher | 24,244 (18%) | 19,716 (17%) | 4,829 (12%) | 3,951 (12%) | 5,111 (15%) | 3,397 (11%) | 4,737 (16%) | 3,706 (14%) | 9,567 (36%) | 8,662 (34%) |
| Borough | | | | | | | | | | |
| Bronx | 30,032 (21%) | 24,529 (20%) | 3,747 (7.5%) | 2,699 (7.4%) | 12,235 (32%) | 9,352 (29%) | 3,444 (12%) | 2,957 (11%) | 10,606 (39%) | 9,521 (37%) |
| Brooklyn | 40,471 (28%) | 34,471 (29%) | 13,812 (28%) | 11,117 (31%) | 3,416 (8.9%) | 2,984 (9.3%) | 16,867 (57%) | 14,313 (55%) | 6,376 (24%) | 6,057 (23%) |
| Manhattan | 22,771 (16%) | 19,151 (16%) | 14,599 (29%) | 11,755 (32%) | 2,401 (6.3%) | 2,128 (6.6%) | 2,105 (7.1%) | 1,889 (7.3%) | 3,666 (14%) | 3,379 (13%) |
| Queens | 30,132 (21%) | 29,354 (24%) | 4,741 (9.5%) | 3,872 (11%) | 13,597 (35%) | 12,724 (40%) | 6,258 (21%) | 6,243 (24%) | 5,536 (21%) | 6,515 (25%) |
| Staten Island | 8,917 (6.1%) | 6,408 (5.3%) | 5,207 (10%) | 3,411 (9.4%) | 3,229 (8.4%) | 2,722 (8.5%) | 405 (1.4%) | 234 (0.9%) | 76 (0.3%) | 41 (0.2%) |
| Non-NYC | 12,731 (8.8%) | 6,362 (5.3%) | 7,871 (16%) | 3,403 (9.4%) | 3,482 (9.1%) | 2,152 (6.7%) | 767 (2.6%) | 384 (1.5%) | 611 (2.3%) | 423 (1.6%) |

Notes:

Abbreviations: NYC = New York City; SOI = severity of illness; NH = non-Hispanic

[a] Hospital quartiles are defined based on a hospital's 2017-2019 percentage of adult hospitalizations where the primary payer was Medicaid or self-pay. Hierarchical algorithm based on common payment logic was used to identify primary payer.

[b] Excludes specialty hospitals and hospitals without ICUs (N=6).

[c] Pre-pandemic period is defined as CDC weeks 2-10 of 2020, corresponding to January 5-March 7, 2020 (based on date of admission).

[d] Wave 1 of COVID-19 is defined as CDC weeks 11-19 of 2020, corresponding to March 8-May 9, 2020 (based on date of admission).

[e] Severity of illness is based on All Patient Refined Diagnosis Related Groups (APR-DRG) classification.

Source: New York State Department of Health, Statewide Planning and Research Cooperative System (SPARCS) 2020 inpatient data, 2022 release.

Table S2: Patient and admission characteristics of ICU hospitalizations among adults (18+) in NYC by hospital quartile, January 5-May 9, 2020[a, b]

| Characteristic, No. (%) | NYC (N=47 hospitals)[c] | | Quartile 1 (lowest % of Medicaid or Self-pay patients) (N=11 hospitals) | | Quartile 2 (N=12 hospitals) | | Quartile 3 (N=12 hospitals) | | Quartile 4 (highest % of Medicaid or Self-pay patients) (N=12 hospitals) | |
|---|---|---|---|---|---|---|---|---|---|---|
| | **Pre-pandemic[d]** | **Wave 1[e]** | **Pre-pandemic** | **Wave 1** | **Pre-pandemic** | **Wave 1** | **Pre-pandemic** | **Wave 1** | **Pre-pandemic** | **Wave 1** |
| ICU hospitalizations | 16,108 (100%) | 15,948 (100%) | 5,986 (100%) | 5,015 (100%) | 3,799 (100%) | 3,680 (100%) | 3,484 (100%) | 3,770 (100%) | 2,839 (100%) | 3,483 (100%) |
| Elixhauser comorbidity score, mean (SD) | 12.7 (10.7) | 12.4 (9.6) | 13.0 (10.9) | 13.7 (9.9) | 13.2 (10.5) | 12.5 (9.7) | 12.2 (10.6) | 11.6 (9.3) | 12.0 (10.7) | 11.5 (9.2) |
| Nursing home admission | 1,011 (6.3%) | 1,019 (6.4%) | 181 (3.0%) | 175 (3.5%) | 308 (8.1%) | 241 (6.5%) | 372 (11%) | 456 (12%) | 150 (5.3%) | 147 (4.2%) |
| Emergency department admission | 11,294 (70%) | 13,866 (87%) | 3,317 (55%) | 3,962 (79%) | 2,546 (67%) | 3,246 (88%) | 2,986 (86%) | 3,576 (95%) | 2,445 (86%) | 3,082 (88%) |
| Major SOI present on admission | 5,188 (32%) | 4,380 (27%) | 1,898 (32%) | 1,389 (28%) | 1,260 (33%) | 1,066 (29%) | 1,047 (30%) | 1,033 (27%) | 983 (35%) | 892 (26%) |
| Extreme SOI present on admission[f] | 4,547 (28%) | 5,226 (33%) | 1,484 (25%) | 1,549 (31%) | 1,089 (29%) | 1,157 (31%) | 1,066 (31%) | 1,218 (32%) | 908 (32%) | 1,302 (37%) |
| Gender | | | | | | | | | | |
| Female | 7,059 (44%) | 6,154 (39%) | 2,572 (43%) | 1,962 (39%) | 1,666 (44%) | 1,484 (40%) | 1,608 (46%) | 1,434 (38%) | 1,213 (43%) | 1,274 (37%) |
| Male | 9,047 (56%) | 9,793 (61%) | 3,413 (57%) | 3,053 (61%) | 2,133 (56%) | 2,196 (60%) | 1,876 (54%) | 2,336 (62%) | 1,625 (57%) | 2,208 (63%) |
| Age group | | | | | | | | | | |
| Age 18-39 | 2,035 (13%) | 1,604 (10%) | 767 (13%) | 506 (10%) | 396 (10%) | 343 (9.3%) | 424 (12%) | 346 (9.2%) | 448 (16%) | 409 (12%) |
| Age 40-64 | 6,146 (38%) | 6,719 (42%) | 2,178 (36%) | 1,917 (38%) | 1,415 (37%) | 1,651 (45%) | 1,258 (36%) | 1,502 (40%) | 1,295 (46%) | 1,649 (47%) |
| Age 65-79 | 5,109 (32%) | 5,518 (35%) | 2,016 (34%) | 1,862 (37%) | 1,272 (33%) | 1,248 (34%) | 1,065 (31%) | 1,323 (35%) | 756 (27%) | 1,085 (31%) |
| Age 80+ | 2,818 (17%) | 2,107 (13%) | 1,025 (17%) | 730 (15%) | 716 (19%) | 438 (12%) | 737 (21%) | 599 (16%) | 340 (12%) | 340 (9.8%) |
| Race/ethnicity | | | | | | | | | | |
| Hispanic | 3,436 (21%) | 4,362 (27%) | 882 (15%) | 999 (20%) | 827 (22%) | 1,056 (29%) | 665 (19%) | 911 (24%) | 1,062 (37%) | 1,396 (40%) |
| NH Asian or Pacific Islander | 1,105 (6.9%) | 1,130 (7.1%) | 362 (6.0%) | 346 (6.9%) | 337 (8.9%) | 287 (7.8%) | 288 (8.3%) | 353 (9.4%) | 118 (4.2%) | 144 (4.1%) |
| NH Black | 4,237 (26%) | 4,165 (26%) | 974 (16%) | 825 (16%) | 1,072 (28%) | 1,113 (30%) | 1,107 (32%) | 1,062 (28%) | 1,084 (38%) | 1,165 (33%) |
| NH White | 4,853 (30%) | 3,632 (23%) | 2,680 (45%) | 1,882 (38%) | 1,021 (27%) | 664 (18%) | 959 (28%) | 919 (24%) | 193 (6.8%) | 167 (4.8%) |
| Other | 2,477 (15%) | 2,659 (17%) | 1,088 (18%) | 963 (19%) | 542 (14%) | 560 (15%) | 465 (13%) | 525 (14%) | 382 (13%) | 611 (18%) |
| Insurance | | | | | | | | | | |
| Medicaid | 4,401 (27%) | 4,708 (30%) | 1,101 (18%) | 1,070 (21%) | 933 (25%) | 1,004 (27%) | 1,071 (31%) | 1,225 (32%) | 1,296 (46%) | 1,409 (40%) |
| Medicare | 8,450 (52%) | 7,935 (50%) | 3,250 (54%) | 2,712 (54%) | 2,175 (57%) | 1,787 (49%) | 1,907 (55%) | 1,981 (53%) | 1,118 (39%) | 1,455 (42%) |
| Private | 2,835 (18%) | 2,971 (19%) | 1,480 (25%) | 1,135 (23%) | 643 (17%) | 832 (23%) | 425 (12%) | 492 (13%) | 287 (10%) | 512 (15%) |
| Self-pay | 286 (1.8%) | 246 (1.5%) | 86 (1.4%) | 57 (1.1%) | 35 (0.9%) | 37 (1.0%) | 54 (1.5%) | 59 (1.6%) | 111 (3.9%) | 93 (2.7%) |
| Other | 136 (0.8%) | 88 (0.6%) | 69 (1.2%) | 41 (0.8%) | 13 (0.3%) | 20 (0.5%) | 27 (0.8%) | 13 (0.3%) | 27 (1.0%) | 14 (0.4%) |
| Neighborhood poverty | | | | | | | | | | |
| 0 to <10% | 2,649 (19%) | 2,278 (15%) | 1,435 (31%) | 1,197 (27%) | 734 (23%) | 611 (18%) | 290 (8.5%) | 270 (7.3%) | 190 (6.9%) | 200 (5.8%) |
| 10 to <20% | 5,898 (42%) | 6,723 (45%) | 1,798 (39%) | 1,813 (41%) | 1,479 (46%) | 1,767 (52%) | 1,738 (51%) | 1,936 (52%) | 883 (32%) | 1,207 (35%) |
| 20 to <30% | 3,164 (23%) | 3,427 (23%) | 946 (21%) | 984 (22%) | 610 (19%) | 654 (19%) | 999 (29%) | 1,092 (30%) | 609 (22%) | 697 (20%) |

| | | | | | | | | | | |
|---|---|---|---|---|---|---|---|---|---|---|
| 30% or higher | 2,305 (16%) | 2,559 (17%) | 431 (9.3%) | 480 (11%) | 413 (13%) | 354 (10%) | 374 (11%) | 401 (11%) | 1,087 (39%) | 1,324 (39%) |
| Borough | | | | | | | | | | |
| Bronx | 2,678 (17%) | 2,909 (18%) | 321 (5.4%) | 293 (5.8%) | 958 (25%) | 992 (27%) | 236 (6.8%) | 310 (8.2%) | 1,163 (41%) | 1,314 (38%) |
| Brooklyn | 4,655 (29%) | 4,843 (30%) | 1,391 (23%) | 1,361 (27%) | 464 (12%) | 458 (12%) | 2,158 (62%) | 2,145 (57%) | 642 (23%) | 879 (25%) |
| Manhattan | 2,466 (15%) | 2,607 (16%) | 1,509 (25%) | 1,539 (31%) | 306 (8.1%) | 340 (9.2%) | 221 (6.3%) | 249 (6.6%) | 430 (15%) | 479 (14%) |
| Queens | 3,183 (20%) | 3,761 (24%) | 652 (11%) | 688 (14%) | 1,241 (33%) | 1,328 (36%) | 761 (22%) | 986 (26%) | 529 (19%) | 759 (22%) |
| Staten Island | 1,083 (6.7%) | 915 (5.7%) | 759 (13%) | 610 (12%) | 290 (7.6%) | 282 (7.7%) | 28 (0.8%) | 21 (0.6%) | * | * |
| Non-NYC | 2,043 (13%) | 913 (5.7%) | 1,354 (23%) | 524 (10%) | 540 (14%) | 280 (7.6%) | 80 (2.3%) | 59 (1.6%) | 69 (2.4%) | 50 (1.4%) |

Notes:

Abbreviations: ICU = intensive care unit; NYC = New York City; SOI = severity of illness; NH = non-Hispanic

[a] ICU use is defined based on revenue codes 200, 201, 202, 203,204, 207, 208, 209, 210, 211, 212, 213 and 219 (Weissman et al. 2017).

[b] Hospital quartiles are defined based on a hospital's 2017-2019 percentage of adult hospitalizations where the primary payer was Medicaid or self-pay. Hierarchical algorithm based on common payment logic was used to identify primary payer.

[c] Excludes specialty hospitals and hospitals without ICUs (N=6).

[d] Pre-pandemic period is defined as CDC weeks 2-10 of 2020, corresponding to January 5-March 7, 2020 (based on date of admission).

[e] Wave 1 of COVID-19 is defined as CDC weeks 11-19 of 2020, corresponding to March 8-May 9, 2020 (based on date of admission).

[f] Severity of illness is based on All Patient Refined Diagnosis Related Groups (APR-DRG) classification.

[g] * indicates suppression of data due to count < 10.

Source: New York State Department of Health, Statewide Planning and Research Cooperative System (SPARCS) 2020 inpatient data, 2022 release.

Table S3: Patient and admission characteristics of non-ICU hospitalizations among adults (18+) in NYC by hospital quartile, January 5-May 9, 2020[a, b]

| **Characteristic, No. (%)** | **NYC** (N=47 hospitals)[c] | | **Quartile 1 (lowest % of Medicaid or Self-pay patients)** (N=11 hospitals) | | **Quartile 2** (N=12 hospitals) | | **Quartile 3** (N=12 hospitals) | | **Quartile 4 (highest % of Medicaid or Self-pay patients)** (N=12 hospitals) | |
|---|---|---|---|---|---|---|---|---|---|---|
| | **Pre-pandemic**[d] | **Wave 1**[e] | **Pre-pandemic** | **Wave 1** | **Pre-pandemic** | **Wave 1** | **Pre-pandemic** | **Wave 1** | **Pre-pandemic** | **Wave 1** |
| Non-ICU hospitalizations | 128,946 (100%) | 104,327 (100%) | 43,991 (100%) | 31,242 (100%) | 34,561 (100%) | 28,382 (100%) | 26,362 (100%) | 22,250 (100%) | 24,032 (100%) | 22,453 (100%) |
| Elixhauser comorbidity score, mean (SD) | 5.5 (8.8) | 5.8 (8.4) | 6.0 (8.8) | 6.0 (8.5) | 6.4 (8.9) | 6.6 (8.5) | 5.1 (8.7) | 5.7 (8.2) | 3.9 (8.2) | 4.6 (8.0) |
| Nursing home admission | 5,193 (4.0%) | 7,018 (6.7%) | 1,051 (2.4%) | 1,170 (3.7%) | 2,141 (6.2%) | 2,825 (10.0%) | 1,637 (6.2%) | 2,348 (11%) | 364 (1.5%) | 675 (3.0%) |
| Emergency department | 89,310 (69%) | 82,087 (79%) | 26,747 (61%) | 22,720 (73%) | 23,347 (68%) | 22,787 (80%) | 20,409 (77%) | 18,634 (84%) | 18,807 (78%) | 17,946 (80%) |
| Major SOI present on admission[f] | 25,180 (20%) | 20,677 (20%) | 8,943 (20%) | 6,610 (21%) | 7,357 (21%) | 5,956 (21%) | 4,815 (18%) | 4,424 (20%) | 4,065 (17%) | 3,687 (16%) |
| Extreme SOI present on admission | 4,865 (3.8%) | 6,728 (6.4%) | 1,744 (4.0%) | 1,847 (5.9%) | 1,660 (4.8%) | 2,227 (7.8%) | 966 (3.7%) | 1,587 (7.1%) | 495 (2.1%) | 1,067 (4.8%) |
| Gender | | | | | | | | | | |
| Female | 73,725 (57%) | 55,518 (53%) | 26,249 (60%) | 18,287 (59%) | 20,342 (59%) | 15,149 (53%) | 14,809 (56%) | 11,617 (52%) | 12,325 (51%) | 10,465 (47%) |
| Male | 55,208 (43%) | 48,798 (47%) | 17,740 (40%) | 12,948 (41%) | 14,219 (41%) | 13,233 (47%) | 11,552 (44%) | 10,633 (48%) | 11,697 (49%) | 11,984 (53%) |
| Age group | | | | | | | | | | |
| Age 18-39 | 36,639 (28%) | 28,158 (27%) | 12,392 (28%) | 9,426 (30%) | 8,154 (24%) | 6,509 (23%) | 7,965 (30%) | 5,952 (27%) | 8,128 (34%) | 6,271 (28%) |
| Age 40-64 | 44,016 (34%) | 35,243 (34%) | 13,625 (31%) | 9,488 (30%) | 11,476 (33%) | 9,574 (34%) | 8,916 (34%) | 7,151 (32%) | 9,999 (42%) | 9,030 (40%) |
| Age 65-79 | 29,363 (23%) | 25,180 (24%) | 10,924 (25%) | 7,455 (24%) | 8,909 (26%) | 7,532 (27%) | 5,416 (21%) | 5,330 (24%) | 4,114 (17%) | 4,863 (22%) |
| Age 80+ | 18,928 (15%) | 15,746 (15%) | 7,050 (16%) | 4,873 (16%) | 6,022 (17%) | 4,767 (17%) | 4,065 (15%) | 3,817 (17%) | 1,791 (7.5%) | 2,289 (10%) |
| Race/ethnicity | | | | | | | | | | |
| Hispanic | 31,595 (25%) | 27,665 (27%) | 7,392 (17%) | 5,918 (19%) | 8,758 (25%) | 7,779 (27%) | 6,533 (25%) | 5,800 (26%) | 8,912 (37%) | 8,168 (36%) |
| NH Asian or Pacific Islander | 9,208 (7.1%) | 7,658 (7.3%) | 2,681 (6.1%) | 2,079 (6.7%) | 3,144 (9.1%) | 2,575 (9.1%) | 2,381 (9.0%) | 2,061 (9.3%) | 1,002 (4.2%) | 943 (4.2%) |
| NH Black | 35,292 (27%) | 28,856 (28%) | 8,504 (19%) | 6,379 (20%) | 10,400 (30%) | 8,456 (30%) | 6,933 (26%) | 5,511 (25%) | 9,455 (39%) | 8,510 (38%) |
| NH White | 37,174 (29%) | 26,306 (25%) | 19,861 (45%) | 12,681 (41%) | 8,316 (24%) | 6,064 (21%) | 7,357 (28%) | 6,048 (27%) | 1,640 (6.8%) | 1,513 (6.7%) |
| Other | 15,677 (12%) | 13,842 (13%) | 5,553 (13%) | 4,185 (13%) | 3,943 (11%) | 3,508 (12%) | 3,158 (12%) | 2,830 (13%) | 3,023 (13%) | 3,319 (15%) |
| Insurance | | | | | | | | | | |
| Medicaid | 45,901 (36%) | 36,484 (35%) | 9,279 (21%) | 7,177 (23%) | 10,561 (31%) | 8,803 (31%) | 12,120 (46%) | 9,331 (42%) | 13,941 (58%) | 11,173 (50%) |
| Medicare | 52,577 (41%) | 43,138 (41%) | 19,432 (44%) | 13,057 (42%) | 16,382 (47%) | 12,944 (46%) | 10,306 (39%) | 9,591 (43%) | 6,457 (27%) | 7,546 (34%) |
| Private | 26,761 (21%) | 22,246 (21%) | 13,934 (32%) | 10,230 (33%) | 7,125 (21%) | 6,231 (22%) | 3,268 (12%) | 2,841 (13%) | 2,434 (10%) | 2,944 (13%) |
| Self-pay | 2,471 (1.9%) | 1,804 (1.7%) | 668 (1.5%) | 479 (1.5%) | 268 (0.8%) | 203 (0.7%) | 447 (1.7%) | 391 (1.8%) | 1,088 (4.5%) | 731 (3.3%) |
| Other | 1,236 (1.0%) | 655 (0.6%) | 678 (1.5%) | 299 (1.0%) | 225 (0.7%) | 201 (0.7%) | 221 (0.8%) | 96 (0.4%) | 112 (0.5%) | 59 (0.3%) |
| Neighborhood poverty | | | | | | | | | | |
| 0 to <10% | 20,455 (17%) | 15,690 (16%) | 9,948 (27%) | 6,883 (24%) | 6,685 (21%) | 5,445 (21%) | 2,056 (8.0%) | 1,692 (7.7%) | 1,766 (7.5%) | 1,670 (7.6%) |
| 10 to <20% | 48,806 (41%) | 43,768 (44%) | 14,991 (40%) | 11,604 (41%) | 14,072 (45%) | 13,147 (50%) | 11,715 (46%) | 10,390 (47%) | 8,028 (34%) | 8,627 (39%) |
| 20 to <30% | 26,566 (23%) | 21,905 (22%) | 8,031 (21%) | 6,316 (22%) | 5,851 (19%) | 4,662 (18%) | 7,484 (29%) | 6,499 (30%) | 5,200 (22%) | 4,428 (20%) |

| | | | | | | | | | | |
|---|---|---|---|---|---|---|---|---|---|---|
| 30% or higher | 21,939 (19%) | 17,157 (17%) | 4,398 (12%) | 3,471 (12%) | 4,698 (15%) | 3,043 (12%) | 4,363 (17%) | 3,305 (15%) | 8,480 (36%) | 7,338 (33%) |
| Borough | | | | | | | | | | |
| Bronx | 27,354 (21%) | 21,620 (21%) | 3,426 (7.8%) | 2,406 (7.7%) | 11,277 (33%) | 8,360 (29%) | 3,208 (12%) | 2,647 (12%) | 9,443 (39%) | 8,207 (37%) |
| Brooklyn | 35,816 (28%) | 29,628 (28%) | 12,421 (28%) | 9,756 (31%) | 2,952 (8.5%) | 2,526 (8.9%) | 14,709 (56%) | 12,168 (55%) | 5,734 (24%) | 5,178 (23%) |
| Manhattan | 20,305 (16%) | 16,544 (16%) | 13,090 (30%) | 10,216 (33%) | 2,095 (6.1%) | 1,788 (6.3%) | 1,884 (7.1%) | 1,640 (7.4%) | 3,236 (13%) | 2,900 (13%) |
| Queens | 26,949 (21%) | 25,593 (25%) | 4,089 (9.3%) | 3,184 (10%) | 12,356 (36%) | 11,396 (40%) | 5,497 (21%) | 5,257 (24%) | 5,007 (21%) | 5,756 (26%) |
| Staten Island | 7,834 (6.1%) | 5,493 (5.3%) | 4,448 (10%) | 2,801 (9.0%) | 2,939 (8.5%) | 2,440 (8.6%) | 377 (1.4%) | 213 (1.0%) | 70 (0.3%) | 39 (0.2%) |
| Non-NYC | 10,688 (8.3%) | 5,449 (5.2%) | 6,517 (15%) | 2,879 (9.2%) | 2,942 (8.5%) | 1,872 (6.6%) | 687 (2.6%) | 325 (1.5%) | 542 (2.3%) | 373 (1.7%) |

Notes:

Abbreviations: ICU = intensive care unit; NYC = New York City; SOI = severity of illness; NH = non-Hispanic

[a] ICU use is defined based on revenue codes 200, 201, 202, 203,204, 207, 208, 209, 210, 211, 212, 213 and 219 (Weissman et al. 2017). Non-ICU use is defined as any admissions without revenue codes for ICU use but may include intermediate ICU use.

[b] Hospital quartiles are defined based on a hospital's 2017-2019 percentage of adult hospitalizations where the primary payer was Medicaid or self-pay. Hierarchical algorithm based on common payment logic was used to identify primary payer.

[c] Excludes specialty hospitals and hospitals without ICUs (N=6).

[d] Pre-pandemic period is defined as CDC weeks 2-10 of 2020, corresponding to January 5-March 7, 2020 (based on date of admission).

[e] Wave 1 of COVID-19 is defined as CDC weeks 11-19 of 2020, corresponding to March 8-May 9, 2020 (based on date of admission).

[f] Severity of illness is based on All Patient Refined Diagnosis Related Groups (APR-DRG) classification.

Source: New York State Department of Health, Statewide Planning and Research Cooperative System (SPARCS) 2020 inpatient data, 2022 release.

Table S4: Patient and admission characteristics of overall mortality among adults (18+) in NYC by hospital quartile, January 5-May 9, 2020[a]

| **Characteristic, No. (%)** | **NYC** (N=47 hospitals)[b] | | **Quartile 1 (lowest % of Medicaid or Self-pay patients)** (N=11 hospitals) | | **Quartile 2** (N=12 hospitals) | | **Quartile 3** (N=12 hospitals) | | **Quartile 4 (highest % of Medicaid or Self-pay patients)** (N=12 hospitals) | |
|---|---|---|---|---|---|---|---|---|---|---|
| | **Pre-pandemic**[c] | **Wave 1**[d] | **Pre-pandemic** | **Wave 1** | **Pre-pandemic** | **Wave 1** | **Pre-pandemic** | **Wave 1** | **Pre-pandemic** | **Wave 1** |
| Deaths among overall admissions | 4,885 (100%) | 18,147 (100%) | 1,602 (100%) | 4,167 (100%) | 1,506 (100%) | 5,263 (100%) | 1,173 (100%) | 4,917 (100%) | 604 (100%) | 3,800 (100%) |
| Elixhauser comorbidity score, mean (SD) | 20.9 (10.4) | 13.5 (9.3) | 21.9 (10.6) | 15.5 (9.9) | 21.4 (10.3) | 14.0 (9.2) | 19.4 (10.1) | 12.2 (8.8) | 20.2 (10.6) | 12.1 (8.8) |
| Nursing home admission | 729 (15%) | 2,924 (16%) | 125 (7.8%) | 435 (10%) | 312 (21%) | 1,102 (21%) | 237 (20%) | 1,074 (22%) | 55 (9.1%) | 313 (8.2%) |
| Emergency department | 4,212 (86%) | 17,082 (94%) | 1,299 (81%) | 3,713 (89%) | 1,307 (87%) | 5,051 (96%) | 1,058 (90%) | 4,765 (97%) | 548 (91%) | 3,553 (94%) |
| Major SOI present on admission[e] | 1,878 (38%) | 4,681 (26%) | 671 (42%) | 1,149 (28%) | 585 (39%) | 1,364 (26%) | 417 (36%) | 1,294 (26%) | 205 (34%) | 874 (23%) |
| Extreme SOI present on admission | 2,484 (51%) | 6,032 (33%) | 751 (47%) | 1,483 (36%) | 766 (51%) | 1,725 (33%) | 615 (52%) | 1,553 (32%) | 352 (58%) | 1,271 (33%) |
| Gender | | | | | | | | | | |
| Female | 2,481 (51%) | 7,443 (41%) | 796 (50%) | 1,769 (42%) | 788 (52%) | 2,241 (43%) | 598 (51%) | 1,991 (40%) | 299 (50%) | 1,442 (38%) |
| Male | 2,404 (49%) | 10,704 (59%) | 806 (50%) | 2,398 (58%) | 718 (48%) | 3,022 (57%) | 575 (49%) | 2,926 (60%) | 305 (50%) | 2,358 (62%) |
| Age group | | | | | | | | | | |
| Age 18-39 | 145 (3.0%) | 424 (2.3%) | 71 (4.4%) | 88 (2.1%) | 29 (1.9%) | 115 (2.2%) | 20 (1.7%) | 90 (1.8%) | 25 (4.1%) | 131 (3.4%) |
| Age 40-64 | 1,079 (22%) | 4,516 (25%) | 354 (22%) | 840 (20%) | 286 (19%) | 1,262 (24%) | 231 (20%) | 1,187 (24%) | 208 (34%) | 1,227 (32%) |
| Age 65-79 | 1,605 (33%) | 7,062 (39%) | 549 (34%) | 1,600 (38%) | 483 (32%) | 2,062 (39%) | 362 (31%) | 1,934 (39%) | 211 (35%) | 1,466 (39%) |
| Age 80+ | 2,056 (42%) | 6,145 (34%) | 628 (39%) | 1,639 (39%) | 708 (47%) | 1,824 (35%) | 560 (48%) | 1,706 (35%) | 160 (26%) | 976 (26%) |
| Race/ethnicity | | | | | | | | | | |
| Hispanic | 992 (20%) | 4,731 (26%) | 256 (16%) | 820 (20%) | 333 (22%) | 1,292 (25%) | 213 (18%) | 1,181 (24%) | 190 (31%) | 1,438 (38%) |
| NH Asian or Pacific Islander | 400 (8.2%) | 1,318 (7.3%) | 99 (6.2%) | 243 (5.8%) | 139 (9.2%) | 445 (8.5%) | 131 (11%) | 438 (8.9%) | 31 (5.1%) | 192 (5.1%) |
| NH Black | 1,289 (26%) | 4,909 (27%) | 320 (20%) | 797 (19%) | 424 (28%) | 1,671 (32%) | 292 (25%) | 1,178 (24%) | 253 (42%) | 1,263 (33%) |
| NH White | 1,591 (33%) | 4,486 (25%) | 706 (44%) | 1,589 (38%) | 445 (30%) | 1,189 (23%) | 397 (34%) | 1,500 (31%) | 43 (7.1%) | 208 (5.5%) |
| Other | 613 (13%) | 2,703 (15%) | 221 (14%) | 718 (17%) | 165 (11%) | 666 (13%) | 140 (12%) | 620 (13%) | 87 (14%) | 699 (18%) |
| Insurance | | | | | | | | | | |
| Medicaid | 779 (16%) | 3,394 (19%) | 212 (13%) | 502 (12%) | 190 (13%) | 870 (17%) | 174 (15%) | 972 (20%) | 203 (34%) | 1,050 (28%) |
| Medicare | 3,657 (75%) | 12,806 (71%) | 1,206 (75%) | 3,152 (76%) | 1,219 (81%) | 3,807 (72%) | 889 (76%) | 3,537 (72%) | 343 (57%) | 2,310 (61%) |
| Private | 369 (7.6%) | 1,661 (9.2%) | 165 (10%) | 477 (11%) | 88 (5.8%) | 503 (9.6%) | 78 (6.6%) | 333 (6.8%) | 38 (6.3%) | 348 (9.2%) |
| Self-pay | 46 (0.9%) | 215 (1.2%) | 15 (0.9%) | 19 (0.5%) | * | 55 (1.0%) | * | 56 (1.1%) | 18 (3.0%) | 85 (2.2%) |
| Other | 34 (0.7%) | 71 (0.4%) | * | 17 (0.4%) | * | 28 (0.5%) | 23 (2.0%) | 19 (0.4%) | * | * |
| Neighborhood poverty | | | | | | | | | | |
| 0 to <10% | 870 (19%) | 2,552 (14%) | 414 (29%) | 946 (24%) | 303 (22%) | 1,001 (20%) | 113 (9.7%) | 363 (7.4%) | 40 (6.7%) | 242 (6.4%) |
| 10 to <20% | 2,028 (45%) | 8,487 (48%) | 518 (37%) | 1,717 (43%) | 704 (51%) | 2,757 (54%) | 584 (50%) | 2,481 (51%) | 222 (37%) | 1,532 (41%) |
| 20 to <30% | 977 (21%) | 3,859 (22%) | 323 (23%) | 909 (23%) | 224 (16%) | 897 (18%) | 310 (27%) | 1,394 (29%) | 120 (20%) | 659 (17%) |
| 30% or higher | 670 (15%) | 2,795 (16%) | 162 (11%) | 399 (10%) | 144 (10%) | 412 (8.1%) | 152 (13%) | 636 (13%) | 212 (36%) | 1,348 (36%) |

| | | | | | | | | | | |
|---|---|---|---|---|---|---|---|---|---|---|
| Borough | | | | | | | | | | |
| Bronx | 967 (20%) | 3,584 (20%) | 114 (7.1%) | 205 (4.9%) | 488 (32%) | 1,436 (27%) | 124 (11%) | 524 (11%) | 241 (40%) | 1,419 (37%) |
| Brooklyn | 1,396 (29%) | 5,810 (32%) | 467 (29%) | 1,516 (36%) | 108 (7.2%) | 724 (14%) | 673 (57%) | 2,682 (55%) | 148 (25%) | 888 (23%) |
| Manhattan | 740 (15%) | 2,391 (13%) | 501 (31%) | 1,424 (34%) | 101 (6.7%) | 347 (6.6%) | 68 (5.8%) | 253 (5.1%) | 70 (12%) | 367 (9.7%) |
| Queens | 1,133 (23%) | 5,068 (28%) | 133 (8.3%) | 421 (10%) | 577 (38%) | 2,125 (40%) | 289 (25%) | 1,410 (29%) | 134 (22%) | 1,112 (29%) |
| Staten Island | 324 (6.6%) | 878 (4.8%) | 206 (13%) | 417 (10%) | 110 (7.3%) | 447 (8.5%) | * | 14 (0.3%) | * | * |
| Non-NYC | 325 (6.7%) | 416 (2.3%) | 181 (11%) | 184 (4.4%) | 122 (8.1%) | 184 (3.5%) | 12 (1.0%) | 34 (0.7%) | 10 (1.7%) | 14 (0.4%) |

Notes:

Abbreviations: NYC = New York City; SOI = severity of illness; NH = non-Hispanic

[a] Hospital quartiles are defined based on a hospital's 2017-2019 percentage of adult hospitalizations where the primary payer was Medicaid or self-pay. Hierarchical algorithm based on common payment logic was used to identify primary payer.

[b] Excludes specialty hospitals and hospitals without ICUs (N=6).

[c] Pre-pandemic period is defined as CDC weeks 2-10 of 2020, corresponding to January 5-March 7, 2020 (based on date of admission).

[d] Wave 1 of COVID-19 is defined as CDC weeks 11-19 of 2020, corresponding to March 8-May 9, 2020 (based on date of admission).

[e] Severity of illness is based on All Patient Refined Diagnosis Related Groups (APR-DRG) classification.

Source: New York State Department of Health, Statewide Planning and Research Cooperative System (SPARCS) 2020 inpatient data, 2022 release.

Table S5: Patient and admission characteristics of ICU mortality among adults (18+) in NYC by hospital quartile, January 5-May 9, 2020[a, b]

| Characteristic, No. (%) | NYC (N=47 hospitals)[c] | | Quartile 1 (lowest % of Medicaid or Self-pay patients) (N=11 hospitals) | | Quartile 2 (N=12 hospitals) | | Quartile 3 (N=12 hospitals) | | Quartile 4 (highest % of Medicaid or Self-pay patients) (N=12 hospitals) | |
|---|---|---|---|---|---|---|---|---|---|---|
| | **Pre-pandemic[d]** | **Wave 1[e]** | **Pre-pandemic** | **Wave 1** | **Pre-pandemic** | **Wave 1** | **Pre-pandemic** | **Wave 1** | **Pre-pandemic** | **Wave 1** |
| Deaths among ICU admissions | 2,261 (100%) | 6,778 (100%) | 743 (100%) | 1,729 (100%) | 560 (100%) | 1,628 (100%) | 565 (100%) | 1,740 (100%) | 393 (100%) | 1,681 (100%) |
| Elixhauser comorbidity score, mean (SD) | 22.0 (10.6) | 14.7 (9.5) | 23.0 (10.6) | 17.1 (9.9) | 22.7 (10.4) | 15.4 (9.7) | 20.5 (10.5) | 13.5 (9.1) | 20.9 (10.9) | 13.0 (8.9) |
| Nursing home admission | 276 (12%) | 534 (7.9%) | 44 (5.9%) | 82 (4.7%) | 90 (16%) | 135 (8.3%) | 105 (19%) | 240 (14%) | 37 (9.4%) | 77 (4.6%) |
| Emergency department | 1,906 (84%) | 6,234 (92%) | 552 (74%) | 1,520 (88%) | 468 (84%) | 1,516 (93%) | 531 (94%) | 1,691 (97%) | 355 (90%) | 1,507 (90%) |
| Major SOI present on admission[f] | 608 (27%) | 1,564 (23%) | 221 (30%) | 387 (22%) | 154 (28%) | 403 (25%) | 129 (23%) | 424 (24%) | 104 (26%) | 350 (21%) |
| Extreme SOI present on admission | 1,509 (67%) | 2,961 (44%) | 474 (64%) | 788 (46%) | 367 (66%) | 663 (41%) | 391 (69%) | 745 (43%) | 277 (70%) | 765 (46%) |
| Gender | | | | | | | | | | |
| Female | 1,008 (45%) | 2,414 (36%) | 317 (43%) | 615 (36%) | 252 (45%) | 609 (37%) | 261 (46%) | 611 (35%) | 178 (45%) | 579 (34%) |
| Male | 1,253 (55%) | 4,364 (64%) | 426 (57%) | 1,114 (64%) | 308 (55%) | 1,019 (63%) | 304 (54%) | 1,129 (65%) | 215 (55%) | 1,102 (66%) |
| Age group | | | | | | | | | | |
| Age 18-39 | 104 (4.6%) | 294 (4.3%) | 48 (6.5%) | 69 (4.0%) | 23 (4.1%) | 68 (4.2%) | 13 (2.3%) | 62 (3.6%) | 20 (5.1%) | 95 (5.7%) |
| Age 40-64 | 625 (28%) | 2,503 (37%) | 194 (26%) | 514 (30%) | 147 (26%) | 640 (39%) | 137 (24%) | 613 (35%) | 147 (37%) | 736 (44%) |
| Age 65-79 | 818 (36%) | 2,806 (41%) | 283 (38%) | 800 (46%) | 199 (36%) | 659 (40%) | 197 (35%) | 713 (41%) | 139 (35%) | 634 (38%) |
| Age 80+ | 714 (32%) | 1,175 (17%) | 218 (29%) | 346 (20%) | 191 (34%) | 261 (16%) | 218 (39%) | 352 (20%) | 87 (22%) | 216 (13%) |
| Race/ethnicity | | | | | | | | | | |
| Hispanic | 455 (20%) | 1,917 (28%) | 102 (14%) | 366 (21%) | 120 (21%) | 424 (26%) | 111 (20%) | 456 (26%) | 122 (31%) | 671 (40%) |
| NH Asian or Pacific Islander | 171 (7.6%) | 530 (7.8%) | 51 (6.9%) | 115 (6.7%) | 44 (7.9%) | 139 (8.5%) | 56 (9.9%) | 189 (11%) | 20 (5.1%) | 87 (5.2%) |
| NH Black | 638 (28%) | 1,750 (26%) | 154 (21%) | 316 (18%) | 166 (30%) | 512 (31%) | 156 (28%) | 394 (23%) | 162 (41%) | 528 (31%) |
| NH White | 641 (28%) | 1,392 (21%) | 309 (42%) | 582 (34%) | 149 (27%) | 295 (18%) | 158 (28%) | 447 (26%) | 25 (6.4%) | 68 (4.0%) |
| Other | 356 (16%) | 1,189 (18%) | 127 (17%) | 350 (20%) | 81 (14%) | 258 (16%) | 84 (15%) | 254 (15%) | 64 (16%) | 327 (19%) |
| Insurance | | | | | | | | | | |
| Medicaid | 449 (20%) | 1,730 (26%) | 114 (15%) | 281 (16%) | 98 (18%) | 369 (23%) | 108 (19%) | 489 (28%) | 129 (33%) | 591 (35%) |
| Medicare | 1,571 (69%) | 3,958 (58%) | 531 (71%) | 1,160 (67%) | 409 (73%) | 931 (57%) | 411 (73%) | 1,048 (60%) | 220 (56%) | 819 (49%) |
| Private | 205 (9.1%) | 960 (14%) | 86 (12%) | 273 (16%) | 49 (8.8%) | 296 (18%) | 39 (6.9%) | 175 (10%) | 31 (7.9%) | 216 (13%) |
| Self-pay | 29 (1.3%) | 106 (1.6%) | *[g] | * | * | 22 (1.4%) | * | 27 (1.6%) | 11 (2.8%) | 51 (3.0%) |
| Other | * | 24 (0.4%) | * | * | * | 10 (0.6%) | * | * | * | * |
| Neighborhood poverty | | | | | | | | | | |
| 0 to <10% | 385 (18%) | 902 (14%) | 192 (30%) | 391 (24%) | 106 (21%) | 287 (19%) | 64 (11%) | 130 (7.6%) | 23 (6.0%) | 94 (5.6%) |
| 10 to <20% | 898 (43%) | 3,108 (47%) | 244 (39%) | 701 (43%) | 252 (50%) | 841 (55%) | 277 (49%) | 930 (54%) | 125 (33%) | 636 (38%) |
| 20 to <30% | 471 (23%) | 1,433 (22%) | 136 (22%) | 356 (22%) | 90 (18%) | 274 (18%) | 155 (28%) | 476 (28%) | 90 (23%) | 327 (20%) |
| 30% or higher | 328 (16%) | 1,101 (17%) | 59 (9.4%) | 166 (10%) | 59 (12%) | 140 (9.1%) | 64 (11%) | 182 (11%) | 146 (38%) | 613 (37%) |

| | | | | | | | | | | |
|---|---|---|---|---|---|---|---|---|---|---|
| Borough | | | | | | | | | | |
| Bronx | 427 (19%) | 1,207 (18%) | 42 (5.7%) | 87 (5.0%) | 163 (29%) | 373 (23%) | 59 (10%) | 154 (8.9%) | 163 (41%) | 593 (35%) |
| Brooklyn | 643 (28%) | 2,167 (32%) | 187 (25%) | 558 (32%) | 62 (11%) | 253 (16%) | 301 (53%) | 926 (53%) | 93 (24%) | 430 (26%) |
| Manhattan | 387 (17%) | 1,013 (15%) | 235 (32%) | 545 (32%) | 49 (8.8%) | 152 (9.3%) | 45 (8.0%) | 112 (6.4%) | 58 (15%) | 204 (12%) |
| Queens | 481 (21%) | 1,838 (27%) | 78 (10%) | 238 (14%) | 181 (32%) | 626 (38%) | 153 (27%) | 528 (30%) | 69 (18%) | 446 (27%) |
| Staten Island | 148 (6.5%) | 341 (5.0%) | 91 (12%) | 193 (11%) | 54 (9.6%) | 144 (8.8%) | * | * | * | * |
| Non-NYC | 175 (7.7%) | 212 (3.1%) | 110 (15%) | 108 (6.2%) | 51 (9.1%) | 80 (4.9%) | * | 16 (0.9%) | * | * |

Notes:

Abbreviations: ICU = intensive care unit; NYC = New York City; SOI = severity of illness; NH = non-Hispanic

[a] ICU use is defined based on revenue codes 200, 201, 202, 203,204, 207, 208, 209, 210, 211, 212, 213 and 219 (Weissman et al. 2017).

[b] Hospital quartiles are defined based on a hospital's 2017-2019 percentage of adult hospitalizations where the primary payer was Medicaid or self-pay. Hierarchical algorithm based on common payment logic was used to identify primary payer.

[c] Excludes specialty hospitals and hospitals without ICUs (N=6).

[d] Pre-pandemic period is defined as CDC weeks 2-10 of 2020, corresponding to January 5-March 7, 2020 (based on date of admission).

[e] Wave 1 of COVID-19 is defined as CDC weeks 11-19 of 2020, corresponding to March 8-May 9, 2020 (based on date of admission).

[f] Severity of illness is based on All Patient Refined Diagnosis Related Groups (APR-DRG) classification.

[g] * indicates suppression of data due to count < 10.

Source: New York State Department of Health, Statewide Planning and Research Cooperative System (SPARCS) 2020 inpatient data, 2022 release.

Table S6: Patient and admission characteristics of non-ICU mortality among adults (18+) in NYC by hospital quartile, January 5-May 9, 2020[a, b]

| Characteristic, No. (%) | NYC (N=47 hospitals)[c] | | Quartile 1 (lowest % of Medicaid or Self-pay patients) (N=11 hospitals) | | Quartile 2 (N=12 hospitals) | | Quartile 3 (N=12 hospitals) | | Quartile 4 (highest % of Medicaid or Self-pay patients) (N=12 hospitals) | |
|---|---|---|---|---|---|---|---|---|---|---|
| | **Pre-pandemic[d]** | **Wave 1[e]** | **Pre-pandemic** | **Wave 1** | **Pre-pandemic** | **Wave 1** | **Pre-pandemic** | **Wave 1** | **Pre-pandemic** | **Wave 1** |
| Non-ICU deaths | 2,624 (100%) | 11,369 (100%) | 859 (100%) | 2,438 (100%) | 946 (100%) | 3,635 (100%) | 608 (100%) | 3,177 (100%) | 211 (100%) | 2,119 (100%) |
| Elixhauser comorbidity score, mean (SD) | 20.1 (10.2) | 12.7 (9.0) | 20.9 (10.4) | 14.4 (9.8) | 20.7 (10.1) | 13.4 (8.9) | 18.4 (9.6) | 11.5 (8.5) | 18.8 (9.9) | 11.3 (8.7) |
| Nursing home admission | 453 (17%) | 2,390 (21%) | 81 (9.4%) | 353 (14%) | 222 (23%) | 967 (27%) | 132 (22%) | 834 (26%) | 18 (8.5%) | 236 (11%) |
| Emergency department | 2,306 (88%) | 10,848 (95%) | 747 (87%) | 2,193 (90%) | 839 (89%) | 3,535 (97%) | 527 (87%) | 3,074 (97%) | 193 (91%) | 2,046 (97%) |
| Major SOI present on admission[f] | 1,270 (48%) | 3,117 (27%) | 450 (52%) | 762 (31%) | 431 (46%) | 961 (26%) | 288 (47%) | 870 (27%) | 101 (48%) | 524 (25%) |
| Extreme SOI present on admission | 975 (37%) | 3,071 (27%) | 277 (32%) | 695 (29%) | 399 (42%) | 1,062 (29%) | 224 (37%) | 808 (25%) | 75 (36%) | 506 (24%) |
| Gender | | | | | | | | | | |
| Female | 1,473 (56%) | 5,029 (44%) | 479 (56%) | 1,154 (47%) | 536 (57%) | 1,632 (45%) | 337 (55%) | 1,380 (43%) | 121 (57%) | 863 (41%) |
| Male | 1,151 (44%) | 6,340 (56%) | 380 (44%) | 1,284 (53%) | 410 (43%) | 2,003 (55%) | 271 (45%) | 1,797 (57%) | 90 (43%) | 1,256 (59%) |
| Age group | | | | | | | | | | |
| Age 18-39 | 41 (1.6%) | 130 (1.1%) | 23 (2.7%) | 19 (0.8%) | 6 (0.6%) | 47 (1.3%) | 7 (1.2%) | 28 (0.9%) | 5 (2.4%) | 36 (1.7%) |
| Age 40-64 | 454 (17%) | 2,013 (18%) | 160 (19%) | 326 (13%) | 139 (15%) | 622 (17%) | 94 (15%) | 574 (18%) | 61 (29%) | 491 (23%) |
| Age 65-79 | 787 (30%) | 4,256 (37%) | 266 (31%) | 800 (33%) | 284 (30%) | 1,403 (39%) | 165 (27%) | 1,221 (38%) | 72 (34%) | 832 (39%) |
| Age 80+ | 1,342 (51%) | 4,970 (44%) | 410 (48%) | 1,293 (53%) | 517 (55%) | 1,563 (43%) | 342 (56%) | 1,354 (43%) | 73 (35%) | 760 (36%) |
| Race/ethnicity | | | | | | | | | | |
| Hispanic | 537 (20%) | 2,814 (25%) | 154 (18%) | 454 (19%) | 213 (23%) | 868 (24%) | 102 (17%) | 725 (23%) | 68 (32%) | 767 (36%) |
| NH Asian or Pacific Islander | 229 (8.7%) | 788 (6.9%) | 48 (5.6%) | 128 (5.3%) | 95 (10%) | 306 (8.4%) | 75 (12%) | 249 (7.8%) | 11 (5.2%) | 105 (5.0%) |
| NH Black | 651 (25%) | 3,159 (28%) | 166 (19%) | 481 (20%) | 258 (27%) | 1,159 (32%) | 136 (22%) | 784 (25%) | 91 (43%) | 735 (35%) |
| NH White | 950 (36%) | 3,094 (27%) | 397 (46%) | 1,007 (41%) | 296 (31%) | 894 (25%) | 239 (39%) | 1,053 (33%) | 18 (8.5%) | 140 (6.6%) |
| Other | 257 (9.8%) | 1,514 (13%) | 94 (11%) | 368 (15%) | 84 (8.9%) | 408 (11%) | 56 (9.2%) | 366 (12%) | 23 (11%) | 372 (18%) |
| Insurance | | | | | | | | | | |
| Medicaid | 330 (13%) | 1,664 (15%) | 98 (11%) | 221 (9.1%) | 92 (9.7%) | 501 (14%) | 66 (11%) | 483 (15%) | 74 (35%) | 459 (22%) |
| Medicare | 2,086 (79%) | 8,848 (78%) | 675 (79%) | 1,992 (82%) | 810 (86%) | 2,876 (79%) | 478 (79%) | 2,489 (78%) | 123 (58%) | 1,491 (70%) |
| Private | 164 (6.2%) | 701 (6.2%) | 79 (9.2%) | 204 (8.4%) | 39 (4.1%) | 207 (5.7%) | 39 (6.4%) | 158 (5.0%) | 7 (3.3%) | 132 (6.2%) |
| Self-pay | 17 (0.6%) | 109 (1.0%) | 6 (0.7%) | 13 (0.5%) | 1 (0.1%) | 33 (0.9%) | 3 (0.5%) | 29 (0.9%) | 7 (3.3%) | 34 (1.6%) |
| Other | 27 (1.0%) | 47 (0.4%) | 1 (0.1%) | 8 (0.3%) | 4 (0.4%) | 18 (0.5%) | 22 (3.6%) | 18 (0.6%) | 0 (0%) | 3 (0.1%) |
| Neighborhood poverty | | | | | | | | | | |
| 0 to <10% | 485 (20%) | 1,650 (15%) | 222 (28%) | 555 (24%) | 197 (23%) | 714 (20%) | 49 (8.2%) | 233 (7.4%) | 17 (8.1%) | 148 (7.0%) |
| 10 to <20% | 1,130 (46%) | 5,379 (48%) | 274 (35%) | 1,016 (43%) | 452 (52%) | 1,916 (54%) | 307 (51%) | 1,551 (49%) | 97 (46%) | 896 (42%) |
| 20 to <30% | 506 (21%) | 2,426 (22%) | 187 (24%) | 553 (23%) | 134 (15%) | 623 (18%) | 155 (26%) | 918 (29%) | 30 (14%) | 332 (16%) |
| 30% or higher | 342 (14%) | 1,694 (15%) | 103 (13%) | 233 (9.9%) | 85 (9.8%) | 272 (7.7%) | 88 (15%) | 454 (14%) | 66 (31%) | 735 (35%) |

| | | | | | | | | | | |
|---|---|---|---|---|---|---|---|---|---|---|
| Borough | | | | | | | | | | |
| Bronx | 540 (21%) | 2,377 (21%) | 72 (8.4%) | 118 (4.8%) | 325 (34%) | 1,063 (29%) | 65 (11%) | 370 (12%) | 78 (37%) | 826 (39%) |
| Brooklyn | 753 (29%) | 3,643 (32%) | 280 (33%) | 958 (39%) | 46 (4.9%) | 471 (13%) | 372 (61%) | 1,756 (55%) | 55 (26%) | 458 (22%) |
| Manhattan | 353 (13%) | 1,378 (12%) | 266 (31%) | 879 (36%) | 52 (5.5%) | 195 (5.4%) | 23 (3.8%) | 141 (4.4%) | 12 (5.7%) | 163 (7.7%) |
| Queens | 652 (25%) | 3,230 (28%) | 55 (6.4%) | 183 (7.5%) | 396 (42%) | 1,499 (41%) | 136 (22%) | 882 (28%) | 65 (31%) | 666 (31%) |
| Staten Island | 176 (6.7%) | 537 (4.7%) | 115 (13%) | 224 (9.2%) | 56 (5.9%) | 303 (8.3%) | 5 (0.8%) | 10 (0.3%) | | |
| Non-NYC | 150 (5.7%) | 204 (1.8%) | 71 (8.3%) | 76 (3.1%) | 71 (7.5%) | 104 (2.9%) | 7 (1.2%) | 18 (0.6%) | 1 (0.5%) | 6 (0.3%) |

Notes:

Abbreviations: ICU = intensive care unit; NYC = New York City; SOI = severity of illness; NH = non-Hispanic

[a] ICU use is defined based on revenue codes 200, 201, 202, 203,204, 207, 208, 209, 210, 211, 212, 213 and 219 (Weissman et al. 2017). Non-ICU use is defined as any admissions without revenue codes for ICU use but may include intermediate ICU use.

[b] Hospital quartiles are defined based on a hospital's 2017-2019 percentage of adult hospitalizations where the primary payer was Medicaid or self-pay. Hierarchical algorithm based on common payment logic was used to identify primary payer.

[c] Excludes specialty hospitals and hospitals without ICUs (N=6).

[d] Pre-pandemic period is defined as CDC weeks 2-10 of 2020, corresponding to January 5-March 7, 2020 (based on date of admission).

[e] Wave 1 of COVID-19 is defined as CDC weeks 11-19 of 2020, corresponding to March 8-May 9, 2020 (based on date of admission).

[f] Severity of illness is based on All Patient Refined Diagnosis Related Groups (APR-DRG) classification.

Source: New York State Department of Health, Statewide Planning and Research Cooperative System (SPARCS) 2020 inpatient data, 2022 release.

Table S7: Hierarchical logistic regression estimates for inpatient mortality among adults (18+) in New York City, January 11- May 9, 2020[a]

| *Outcome* | **ICU death[b]** (N=30,326 hospitalizations.) | | **Non-ICU death[c]** (N=219,729 hospitalizations.) | | **Overall inpatient death** (N=250,101 hospitalizations.) | |
|---|---|---|---|---|---|---|
| | Odds Ratio | 95% CI | Odds Ratio | 95% CI | Odds Ratio | 95% CI |
| **Patient or admission characteristic** | | | | | | |
| Female | 0.82*** | (0.77,0.87) | 0.94** | (0.90,0.98) | 0.88*** | (0.85,0.91) |
| **Age group** | | | | | | |
| Age 18-39 [Reference] | 1 | (1.00,1.00) | 1 | (1.00,1.00) | 1 | (1.00,1.00) |
| Age 40-64 | 1.67*** | (1.47,1.91) | 2.91*** | (2.47,3.42) | 2.02*** | (1.83,2.22) |
| Age 65-79 | 2.66*** | (2.29,3.09) | 5.77*** | (4.88,6.82) | 3.44*** | (3.10,3.81) |
| Age 80+ | 3.79*** | (3.21,4.47) | 12.10*** | (10.21,14.35) | 6.02*** | (5.41,6.69) |
| **Race/ethnicity** | | | | | | |
| NH White [Ref.] | 1 | (1.00,1.00) | 1 | (1.00,1.00) | 1 | (1.00,1.00) |
| Hispanic | 1.05 | (0.95,1.17) | 0.99 | (0.92,1.06) | 1.04 | (0.98,1.10) |
| NH Asian or Pacific Islander | 1.12 | (0.98,1.29) | 0.98 | (0.89,1.08) | 1.05 | (0.97,1.13) |
| NH Black | 1.07 | (0.96,1.19) | 0.88*** | (0.82,0.94) | 0.92** | (0.87,0.97) |
| Other race or ethnicity | 1.29*** | (1.16,1.44) | 1.04 | (0.96,1.12) | 1.17*** | (1.10,1.24) |
| **Insurance** | | | | | | |
| Private insurance [Reference] | 1 | (1.00,1.00) | 1 | (1.00,1.00) | 1 | (1.00,1.00) |
| Medicaid | 1.06 | (0.96,1.18) | 1.10* | (1.01,1.21) | 1.03 | (0.96,1.09) |
| Medicare | 1.09 | (0.98,1.22) | 1.27*** | (1.16,1.40) | 1.12** | (1.05,1.20) |
| Self-pay | 1.39* | (1.07,1.82) | 0.93 | (0.74,1.16) | 1.01 | (0.85,1.18) |
| Other insurance | 1.12 | (0.71,1.77) | 2.29*** | (1.70,3.08) | 1.65*** | (1.29,2.10) |
| Nursing home admission | 0.75*** | (0.66,0.85) | 1.40*** | (1.31,1.50) | 1.13*** | (1.07,1.20) |
| Elixhauser comorbidity score | 1.04*** | (1.04,1.05) | 1.05*** | (1.05,1.05) | 1.06*** | (1.05,1.06) |
| Emergency department | 1.11 | (0.98,1.26) | 1.26*** | (1.13,1.41) | 1.12** | (1.03,1.21) |
| **Severity of disease (SOI)[d]** | | | | | | |
| No major or extreme SOI present on admission [Reference] | 1 | (1.00,1.00) | 1 | (1.00,1.00) | 1 | (1.00,1.00) |
| Major SOI present on admission | 1.85*** | (1.66,2.05) | 2.97*** | (2.79,3.17) | 2.95*** | (2.79,3.11) |
| Extreme SOI present on admission | 3.49*** | (3.12,3.89) | 8.33*** | (7.71,8.99) | 8.29*** | (7.80,8.80) |

| | | | | | | |
|---|---|---|---|---|---|---|
| **Hospital type[e, f]** | | | | | | |
| Private - AMC [Reference] | 1 | (1.00,1.00) | 1 | (1.00,1.00) | 1 | (1.00,1.00) |
| Private – other | 1.31* | (1.04,1.64) | 2.27*** | (1.77,2.92) | 1.95*** | (1.58,2.39) |
| Public | 0.98 | (0.78,1.23) | 1.07 | (0.82,1.38) | 1.09 | (0.89,1.35) |
| **Hospital strain (% of pre-pandemic average)[g]** | | | | | | |
| 0% to 100% [Reference] | 1 | (1.00,1.00) | 1 | (1.00,1.00) | 1 | (1.00,1.00) |
| 101% to 150% | 1.17*** | (1.10,1.26) | 1.28*** | (1.22,1.34) | 1.28*** | (1.24,1.34) |
| 151% to 200% | 2.63*** | (2.31,3.00) | 2.60*** | (2.40,2.82) | 2.62*** | (2.45,2.80) |
| > 200% | 3.26*** | (2.82,3.78) | 3.44*** | (3.13,3.78) | 3.36*** | (3.11,3.63) |
| cons[hospital] | 1.09*** | (1.04,1.14) | 1.13*** | (1.07,1.19) | 1.09*** | (1.05,1.13) |
| Principal diagnosis[h] | Yes | | Yes | | Yes | |
| Neighborhood of residence[i] | Yes | | Yes | | Yes | |

Notes:
* p<0.05, ** p<0.01, *** p<0.001
Abbreviations: ICU = intensive care unit; NH = non-Hispanic; SOI = severity of illness; AMC = academic medical center (including affiliates).
[a] < 100 observations were excluded due to missing data.
[b] ICU use is identified based on revenue codes 200, 201, 202, 203,204, 207, 208, 209, 210, 211, 212, 213 and 219 (Weissman et al. 2017).
[c] Non-ICU use is defined as any admissions without revenue codes for ICU use but may include intermediate ICU use.
[d] Severity of illness (SOI) is based on All Patient Refined Diagnosis Related Groups (APR-DRG) classification.
[e] Hospital type is defined as public if a hospital belonged to the NYC H+H system or was state-owned; private (other) hospitals are defined as private hospitals not part of a private AMC system.
[f] Excludes specialty hospitals and hospitals without ICUs (N=6).
[g] Hospital strain is defined as the total number of adult ICU admissions to the hospital in the week prior to and including the patient admission date as a percentage of the hospital's pre-pandemic baseline weekly average ICU admissions.
[h] Principal diagnosis is based on Clinical Classification Software Refined (CCSR) version 2021.1 groupings of ICD-10 diagnostic codes. Top 100 principal diagnoses for hospitalizations resulting in in-hospital death are included. COVID-19 ICD-10 diagnostic code (U.071) went into effect for discharges beginning on April 1, 2020, and does not capture admissions with COVID-19 that resulted in discharges prior to April 1.
[i] Neighborhoods in New York City are defined as Neighborhood Tabulation Areas based on geocoded patient address; separate indicator is included for non-NYC addresses.
Source: New York State Department of Health, Statewide Planning and Research Cooperative System (SPARCS) 2020 inpatient data, 2022 release.

Table S8: Hierarchical logistic regression estimates (with alternative measure 1 of hospital strain) for inpatient mortality among adults in NYC, January 11- May 9, 2020[a, b]

| *Outcome* | **ICU death[c]** (N=30,326 hosp.) | | **Non-ICU death[d]** (N=219,729 hosp.) | | **Overall hospital death** (N=250,101 hosp.) | |
|---|---|---|---|---|---|---|
| | Odds Ratio | CI 95% | Odds Ratio | CI 95% | Odds Ratio | CI 95% |
| **Patient or admission characteristic** | | | | | | |
| Female | 0.82*** | [0.77,0.87] | 0.93** | [0.89,0.97] | 0.88*** | [0.85,0.91] |
| **Age group** | | | | | | |
| Age 18-39 [Reference] | 1 | [1.00,1.00] | 1 | [1.00,1.00] | 1 | [1.00,1.00] |
| Age 40-64 | 1.70*** | [1.49,1.94] | 2.91*** | [2.47,3.42] | 2.03*** | [1.85,2.23] |
| Age 65 79 | 2.72*** | [2.35,3.16] | 5.76*** | [4.87,6.82] | 3.46*** | [3.13,3.84] |
| Age 80+ | 3.81*** | [3.23,4.49] | 12.04*** | [10.16,14.27] | 6.04*** | [5.43,6.71] |
| **Race/ethnicity** | | | | | | |
| NH White [Reference] | 1 | [1.00,1.00] | 1 | [1.00,1.00] | 1 | [1.00,1.00] |
| Hispanic | 1.07 | [0.97,1.19] | 0.99 | [0.92,1.06] | 1.05 | [0.99,1.11] |
| NH Asian or Pacific Islander | 1.12 | [0.97,1.28] | 0.98 | [0.89,1.08] | 1.05 | [0.97,1.13] |
| NH Black | 1.08 | [0.97,1.20] | 0.88*** | [0.82,0.94] | 0.92** | [0.87,0.98] |
| Other race or ethnicity | 1.29*** | [1.16,1.44] | 1.04 | [0.96,1.12] | 1.17*** | [1.10,1.24] |
| **Insurance** | | | | | | |
| Private insurance [Reference] | 1 | [1.00,1.00] | 1 | [1.00,1.00] | 1 | [1.00,1.00] |
| Medicaid | 1.05 | [0.95,1.16] | 1.10* | [1.00,1.20] | 1.02 | [0.95,1.09] |
| Medicare | 1.07 | [0.96,1.20] | 1.26*** | [1.15,1.39] | 1.11** | [1.04,1.19] |
| Self-pay | 1.36* | [1.04,1.77] | 0.93 | [0.74,1.16] | 1 | [0.85,1.17] |
| Other insurance | 1.08 | [0.69,1.71] | 2.19*** | [1.63,2.96] | 1.59*** | [1.25,2.03] |
| Nursing home admission | 0.74*** | [0.65,0.84] | 1.42*** | [1.33,1.52] | 1.14*** | [1.08,1.21] |
| Elixhauser comorbidity score | 1.04*** | [1.04,1.04] | 1.05*** | [1.05,1.05] | 1.05*** | [1.05,1.06] |
| Emergency department | 1.08 | [0.95,1.22] | 1.28*** | [1.15,1.43] | 1.12** | [1.03,1.21] |
| **Severity of illness (SOI)[e]** | | | | | | |
| No major or extreme SOI present on admission [Reference] | 1 | (1.00,1.00) | 1 | (1.00,1.00) | 1 | (1.00,1.00) |
| Major SOI present on admission | 1.86*** | [1.67,2.06] | 2.97*** | [2.79,3.17] | 2.95*** | [2.80,3.11] |

| | | | | | | |
|---|---|---|---|---|---|---|
| Extreme SOI present on admission | 3.48*** | [3.12,3.89] | 8.45*** | [7.83,9.13] | 8.36*** | [7.87,8.88] |
| **Hospital type[f, g]** | | | | | | |
| Private AMC [Reference] | 1 | [1.00,1.00] | 1 | [1.00,1.00] | 1 | [1.00,1.00] |
| Private other | 1.24 | [0.97,1.59] | 2.09*** | [1.56,2.80] | 1.80*** | [1.41,2.30] |
| Public | 1.36* | [1.06,1.75] | 1.62** | [1.20,2.19] | 1.63*** | [1.27,2.09] |
| **Hospital strain (% of certified ICU beds)** | | | | | | |
| 0-25% [Reference] | 1 | [1.00,1.00] | 1 | [1.00,1.00] | 1 | [1.00,1.00] |
| 25-50% | 1.63*** | [1.50,1.78] | 1.74*** | [1.65,1.85] | 1.73*** | [1.65,1.81] |
| 50-75% | 2.01*** | [1.61,2.52] | 2.51*** | [2.13,2.95] | 2.49*** | [2.18,2.84] |
| 75-100% | 3.04*** | [2.22,4.15] | 3.66*** | [2.98,4.49] | 3.63*** | [3.07,4.29] |
| 100% + | 3.06*** | [2.18,4.28] | 6.23*** | [4.92,7.89] | 5.61*** | [4.63,6.79] |
| cons[hospital] | 1.11*** | [1.05,1.17] | 1.18*** | [1.10,1.28] | 1.13*** | [1.07,1.19] |
| Principal diagnosis[h] | Yes | | Yes | | Yes | |
| Neighborhood of residence[i] | Yes | | Yes | | Yes | |

Notes:
* p<0.05, ** p<0.01, *** p<0.001
Abbreviations: ICU = intensive care unit; NH = non-Hispanic; SOI = severity of illness; AMC = academic medical center (including affiliates).
[a] Hospital strain is defined as daily average adult admissions with ICU use over a 7-day period prior to and including patient admission date as a percentage of certified ICU beds.
[b] < 100 observations were excluded due to missing data.
[c] ICU use is identified based on revenue codes 200, 201, 202, 203,204, 207, 208, 209, 210, 211, 212, 213 and 219 (Weissman et al. 2017).
[d] Non-ICU use is defined as any admissions without revenue codes for ICU use but may include intermediate ICU use.
[e] Severity of illness (SOI) is based on All Patient Refined Diagnosis Related Groups (APR-DRG) classification.
[f] Hospital type is defined as public if a hospital belonged to the NYC H+H system or was state-owned; private (other) hospitals are defined as private hospitals not part of a private AMC system.
[g] Excludes specialty hospitals and hospitals without ICUs (N=6).
[h] Principal diagnosis is based on Clinical Classification Software Refined (CCSR) version 2021.1 groupings of ICD-10 diagnostic codes. Top 100 principal diagnoses for hospitalizations resulting in in-hospital death are included. COVID-19 ICD-10 diagnostic code (U.071) went into effect for discharges beginning on April 1, 2020, and does not capture admissions with COVID-19 that resulted in discharges prior to April 1.
[i] Neighborhoods in New York City are defined as Neighborhood Tabulation Areas based on geocoded patient address; separate indicator is included for non-NYC addresses.
Source: New York State Department of Health, Statewide Planning and Research Cooperative System (SPARCS) 2020 inpatient data, 2022 release.

Table S9: Hierarchical logistic regression estimates (with alternative measure 2 of hospital strain) for inpatient mortality among adults (18+) in NYC, January 11- May 9, 2020[a, b]

| *Outcome* | **ICU death**[c] (N=30,326 hosp.) | | **Non-ICU death**[d] (N=219,712 hosp.) | | **Overall hospital death** (N=250,084 hosp.) | |
|---|---|---|---|---|---|---|
| | Odds Ratio | CI 95% | Odds Ratio | CI 95% | Odds Ratio | CI 95% |
| **Patient or admission characteristic** | | | | | | |
| Female | 0.83*** | [0.78,0.88] | 0.97 | [0.93,1.01] | 0.90*** | [0.87,0.93] |
| **Age group** | | | | | | |
| Age 18-39 [Reference] | 1 | [1.00,1.00] | 1 | [1.00,1.00] | 1 | [1.00,1.00] |
| Age 40-64 | 1.68*** | [1.47,1.91] | 2.94*** | [2.50,3.46] | 2.03*** | [1.84,2.23] |
| Age 65 79 | 2.63*** | [2.26,3.06] | 5.66*** | [4.78,6.70] | 3.36*** | [3.03,3.73] |
| Age 80+ | 3.94*** | [3.34,4.65] | 12.06*** | [10.17,14.31] | 5.96*** | [5.36,6.63] |
| **Race/ethnicity** | | | | | | |
| NH White [Reference] | 1 | [1.00,1.00] | 1 | [1.00,1.00] | 1 | [1.00,1.00] |
| Hispanic | 1.02 | [0.91,1.13] | 0.99 | [0.92,1.06] | 1.03 | [0.97,1.09] |
| NH Asian or Pacific Islander | 1.09 | [0.95,1.26] | 1 | [0.91,1.10] | 1.05 | [0.98,1.14] |
| NH Black | 1.05 | [0.94,1.17] | 0.86*** | [0.80,0.92] | 0.90*** | [0.85,0.95] |
| Other race or ethnicity | 1.24*** | [1.11,1.38] | 1.01 | [0.93,1.09] | 1.14*** | [1.07,1.21] |
| **Insurance** | | | | | | |
| Private insurance [Reference] | 1 | [1.00,1.00] | 1 | [1.00,1.00] | 1 | [1.00,1.00] |
| Medicaid | 1.07 | [0.97,1.19] | 1.11* | [1.01,1.22] | 1.03 | [0.97,1.10] |
| Medicare | 1.13* | [1.01,1.27] | 1.33*** | [1.21,1.46] | 1.16*** | [1.08,1.24] |
| Self-pay | 1.52** | [1.17,1.99] | 0.96 | [0.77,1.20] | 1.06 | [0.90,1.24] |
| Other insurance | 1.19 | [0.75,1.88] | 2.26*** | [1.67,3.05] | 1.67*** | [1.31,2.14] |
| Nursing home admission | 0.77*** | [0.68,0.87] | 1.37*** | [1.28,1.46] | 1.11*** | [1.05,1.18] |
| Elixhauser comorbidity score | 1.04*** | [1.04,1.05] | 1.05*** | [1.05,1.06] | 1.06*** | [1.06,1.06] |
| Emergency department | 1.06 | [0.93,1.20] | 1.17** | [1.05,1.31] | 1.05 | [0.97,1.14] |
| **Severity of illness (SOI)**[e] | | | | | | |
| No major or extreme SOI present on admission [Reference] | 1 | (1.00,1.00) | 1 | (1.00,1.00) | 1 | (1.00,1.00) |

| | | | | | | |
|---|---|---|---|---|---|---|
| Major SOI present on admission | 1.78*** | [1.60,1.97] | 2.79*** | [2.61,2.97] | 2.80*** | [2.66,2.96] |
| Extreme SOI present on admission | 3.39*** | [3.04,3.79] | 7.42*** | [6.86,8.02] | 7.85*** | [7.39,8.35] |
| **Hospital type[f, g]** | | | | | | |
| Private AMC [Reference] | 1 | [1.00,1.00] | 1 | [1.00,1.00] | 1 | [1.00,1.00] |
| Private other | 1.45** | [1.15,1.84] | 2.44*** | [1.89,3.16] | 2.11*** | [1.71,2.60] |
| Public | 1.29* | [1.02,1.64] | 1.43** | [1.10,1.86] | 1.49*** | [1.20,1.84] |
| **Hospital strain (% ICU COVID-19 positive)** | | | | | | |
| 0-25% [Reference] | 1 | [1.00,1.00] | 1 | [1.00,1.00] | 1 | [1.00,1.00] |
| 25-50% | 2.21*** | [2.00,2.44] | 2.59*** | [2.40,2.79] | 2.41*** | [2.27,2.56] |
| 50-75% | 2.57*** | [2.33,2.83] | 3.20*** | [2.98,3.44] | 2.79*** | [2.64,2.95] |
| 75-100% | 3.13*** | [2.85,3.43] | 4.21*** | [3.95,4.49] | 3.44*** | [3.27,3.62] |
| cons[hospital] | 1.10*** | [1.05,1.15] | 1.14*** | [1.07,1.21] | 1.09*** | [1.05,1.13] |
| Principal diagnosis[h] | Yes | | Yes | | Yes | |
| Neighborhood of residence[i] | Yes | | Yes | | Yes | |

Notes:
* p<0.05, ** p<0.01, *** p<0.001
Abbreviations: ICU = intensive care unit; NH = non-Hispanic; SOI = severity of illness; AMC = academic medical center (including affiliates).
[a] Hospital strain is defined as adult COVID-19 positive admissions with ICU use over a 7-day period prior to and including patient admission date as a percentage of adult admissions with ICU use.
[b] < 100 observations were excluded due to missing data.
[c] ICU use is identified based on revenue codes 200, 201, 202, 203,204, 207, 208, 209, 210, 211, 212, 213 and 219 (Weissman et al. 2017).
[d] Non-ICU use is defined as any admissions without revenue codes for ICU use but may include intermediate ICU use.
[e] Severity of illness (SOI) is based on All Patient Refined Diagnosis Related Groups (APR-DRG) classification.
[f] Hospital type is defined as public if a hospital belonged to the NYC H+H system or was state-owned; private (other) hospitals are defined as private hospitals not part of a private AMC system.
[g] Excludes specialty hospitals and hospitals without ICUs (N=6).
[h] Principal diagnosis is based on Clinical Classification Software Refined (CCSR) version 2021.1 groupings of ICD-10 diagnostic codes. Top 100 principal diagnoses for hospitalizations resulting in in-hospital death are included. COVID-19 ICD-10 diagnostic code (U.071) went into effect for discharges beginning on April 1, 2020, and does not capture admissions with COVID-19 that resulted in discharges prior to April 1.
[i] Neighborhoods in New York City are defined as Neighborhood Tabulation Areas based on geocoded patient address; separate indicator is included for non-NYC addresses.
Source: New York State Department of Health, Statewide Planning and Research Cooperative System (SPARCS) 2020 inpatient data, 2022 release.

Table S10: Hierarchical logistic regression estimates (with alternative measure 3 of hospital strain) for inpatient mortality among adults (18+) in NYC, January 11- May 9, 2020[a, b]

| *Outcome* | **ICU death[c]** (N=30,326 hosp.) | | **Non-ICU death[d]** (N=219,729 hosp.) | | **Overall hospital death** (N=250,101 hosp.) | |
|---|---|---|---|---|---|---|
| | Odds Ratio | CI 95% | Odds Ratio | CI 95% | Odds Ratio | CI 95% |
| **Patient or admission characteristic** | | | | | | |
| Female | 0.83*** | [0.78,0.89] | 0.96 | [0.92,1.01] | 0.90*** | [0.87,0.93] |
| **Age group** | | | | | | |
| Age 18-39 [Reference] | 1 | [1.00,1.00] | 1 | [1.00,1.00] | 1 | [1.00,1.00] |
| Age 40-64 | 1.65*** | [1.45,1.89] | 2.89*** | [2.46,3.41] | 2.00*** | [1.82,2.20] |
| Age 65 79 | 2.62*** | [2.25,3.05] | 5.69*** | [4.81,6.73] | 3.37*** | [3.04,3.74] |
| Age 80+ | 3.85*** | [3.26,4.55] | 12.26*** | [10.34,14.55] | 6.04*** | [5.43,6.72] |
| **Race/ethnicity** | | | | | | |
| NH White [Reference] | 1 | [1.00,1.00] | 1 | [1.00,1.00] | 1 | [1.00,1.00] |
| Hispanic | 1.02 | [0.92,1.13] | 0.97 | [0.91,1.05] | 1.02 | [0.96,1.08] |
| NH Asian or Pacific Islander | 1.11 | [0.97,1.27] | 1 | [0.91,1.10] | 1.06 | [0.98,1.14] |
| NH Black | 1.04 | [0.93,1.16] | 0.85*** | [0.79,0.91] | 0.89*** | [0.84,0.95] |
| Other race or ethnicity | 1.23*** | [1.11,1.37] | 0.99 | [0.92,1.07] | 1.12*** | [1.06,1.19] |
| **Insurance** | | | | | | |
| Private insurance [Reference] | 1 | [1.00,1.00] | 1 | [1.00,1.00] | 1 | [1.00,1.00] |
| Medicaid | 1.1 | [1.00,1.22] | 1.12* | [1.03,1.23] | 1.05 | [0.99,1.12] |
| Medicare | 1.16** | [1.04,1.30] | 1.32*** | [1.20,1.44] | 1.16*** | [1.09,1.25] |
| Self-pay | 1.47** | [1.13,1.93] | 0.96 | [0.76,1.20] | 1.05 | [0.89,1.23] |
| Other insurance | 1.18 | [0.75,1.87] | 2.25*** | [1.67,3.04] | 1.66*** | [1.30,2.12] |
| Nursing home admission | 0.79*** | [0.69,0.89] | 1.41*** | [1.32,1.51] | 1.14*** | [1.08,1.21] |
| Elixhauser comorbidity score | 1.05*** | [1.04,1.05] | 1.05*** | [1.05,1.06] | 1.06*** | [1.06,1.06] |
| Emergency department | 1.05 | [0.92,1.19] | 1.16** | [1.04,1.30] | 1.04 | [0.96,1.12] |
| **Severity of illness (SOI)[e]** | | | | | | |
| No major or extreme SOI present on admission [Reference] | 1 | (1.00,1.00) | 1 | (1.00,1.00) | 1 | (1.00,1.00) |
| Major SOI | 1.84*** | [1.66,2.05] | 2.98*** | [2.79,3.18] | 2.96*** | [2.80,3.12] |

| | | | | | | |
|---|---|---|---|---|---|---|
| present on admission | | | | | | |
| Extreme SOI present on admission | 3.61*** | [3.23,4.03] | 8.28*** | [7.65,8.95] | 8.47*** | [7.97,9.01] |
| **Hospital type[f, g]** | | | | | | |
| Private AMC [Reference] | 1 | [1.00,1.00] | 1 | [1.00,1.00] | 1 | [1.00,1.00] |
| Private other | 1.52*** | [1.20,1.93] | 2.63*** | [2.01,3.46] | 2.25*** | [1.81,2.81] |
| Public | 1.14 | [0.90,1.45] | 1.23 | [0.93,1.63] | 1.28* | [1.03,1.61] |
| **Hospital strain (sepsis, % of pre-pandemic average)** | | | | | | |
| 0% to <100% [Reference] | 1 | [1.00,1.00] | 1 | [1.00,1.00] | 1 | [1.00,1.00] |
| 100% to <150% | 1.19*** | [1.09,1.29] | 1.19*** | [1.11,1.27] | 1.20*** | [1.14,1.26] |
| 150% to <200% | 2.13*** | [1.90,2.38] | 2.34*** | [2.16,2.54] | 2.27*** | [2.13,2.43] |
| 200% or higher | 3.45*** | [3.13,3.81] | 4.10*** | [3.82,4.40] | 3.71*** | [3.51,3.92] |
| cons[hospital] | 1.10*** | [1.05,1.15] | 1.15*** | [1.08,1.23] | 1.10*** | [1.06,1.15] |
| Principal diagnosis[h] | Yes | | Yes | | Yes | |
| Neighborhood of residence[i] | Yes | | Yes | | Yes | |

Notes:
* p<0.05, ** p<0.01, *** p<0.001
Abbreviations: ICU = intensive care unit; NH = non-Hispanic; SOI = severity of illness; AMC = academic medical center (including affiliates).
[a] Hospital strain is defined as adult sepsis admissions over a 7-day period prior to and including patient admission date in any setting as a percentage of pre-pandemic weekly average adult sepsis admissions.
[b] < 100 observations were excluded due to missing data.
[c] ICU use is identified based on revenue codes 200, 201, 202, 203,204, 207, 208, 209, 210, 211, 212, 213 and 219 (Weissman et al. 2017).
[d] Non-ICU use is defined as any admissions without revenue codes for ICU use but may include intermediate ICU use.
[e] Severity of illness (SOI) is based on All Patient Refined Diagnosis Related Groups (APR-DRG) classification.
[f] Hospital type is defined as public if a hospital belonged to the NYC H+H system or was state-owned; private (other) hospitals are defined as private hospitals not part of a private AMC system.
[g] Excludes specialty hospitals and hospitals without ICUs (N=6).
[h] Principal diagnosis is based on Clinical Classification Software Refined (CCSR) version 2021.1 groupings of ICD-10 diagnostic codes. Top 100 principal diagnoses for hospitalizations resulting in in-hospital death are included. COVID-19 ICD-10 diagnostic code (U.071) went into effect for discharges beginning on April 1, 2020, and does not capture admissions with COVID-19 that resulted in discharges prior to April 1.
[i] Neighborhoods in New York City are defined as Neighborhood Tabulation Areas based on geocoded patient address; separate indicator is included for non-NYC addresses.
Source: New York State Department of Health, Statewide Planning and Research Cooperative System (SPARCS) 2020 inpatient data, 2022 release.

Table S11: Non-hierarchical logistic regression estimates for inpatient mortality among adults (18+) in NYC, January 11- May 9, 2020[a]

| *Outcome* | **ICU death[b]** (N=30,326 hosp.) | | **Non-ICU death[c]** (N=219,729 hosp.) | | **Overall hospital death** (N=250,101 hosp.) | |
|---|---|---|---|---|---|---|
| | Odds Ratio | CI 95% | Odds Ratio | CI 95% | Odds Ratio | CI 95% |
| **Patient or admission characteristic** | | | | | | |
| Female | 0.82*** | [0.77,0.87] | 0.94** | [0.90,0.98] | 0.88*** | [0.85,0.91] |
| **Age group** | | | | | | |
| Age 18-39 [Reference] | 1 | [1.00,1.00] | 1 | [1.00,1.00] | 1 | [1.00,1.00] |
| Age 40-64 | 1.67*** | [1.46,1.90] | 2.91*** | [2.47,3.42] | 2.02*** | [1.83,2.22] |
| Age 65 79 | 2.66*** | [2.29,3.09] | 5.76*** | [4.87,6.81] | 3.43*** | [3.10,3.80] |
| Age 80+ | 3.79*** | [3.21,4.47] | 12.09*** | [10.20,14.33] | 6.01*** | [5.41,6.69] |
| **Race/ethnicity** | | | | | | |
| NH White [Reference] | 1 | [1.00,1.00] | 1 | [1.00,1.00] | 1 | [1.00,1.00] |
| Hispanic | 1.05 | [0.95,1.17] | 0.99 | [0.92,1.06] | 1.04 | [0.98,1.10] |
| NH Asian or Pacific Islander | 1.13 | [0.98,1.29] | 0.98 | [0.89,1.08] | 1.05 | [0.97,1.13] |
| NH Black | 1.07 | [0.96,1.19] | 0.87*** | [0.81,0.94] | 0.91** | [0.86,0.97] |
| Other race or ethnicity | 1.30*** | [1.17,1.44] | 1.04 | [0.96,1.12] | 1.17*** | [1.10,1.25] |
| **Insurance** | | | | | | |
| Private insurance [Reference] | 1 | [1.00,1.00] | 1 | [1.00,1.00] | 1 | [1.00,1.00] |
| Medicaid | 1.06 | [0.96,1.17] | 1.10* | [1.01,1.21] | 1.03 | [0.96,1.09] |
| Medicare | 1.09 | [0.97,1.22] | 1.28*** | [1.16,1.40] | 1.12** | [1.05,1.20] |
| Self-pay | 1.39* | [1.06,1.81] | 0.93 | [0.74,1.16] | 1.01 | [0.85,1.19] |
| Other insurance | 1.1 | [0.70,1.74] | 2.28*** | [1.69,3.07] | 1.64*** | [1.29,2.10] |
| Nursing home admission | 0.74*** | [0.66,0.84] | 1.40*** | [1.31,1.50] | 1.13*** | [1.07,1.20] |
| Elixhauser comorbidity score | 1.04*** | [1.04,1.05] | 1.05*** | [1.05,1.05] | 1.06*** | [1.05,1.06] |
| Emergency department | 1.1 | [0.97,1.25] | 1.24*** | [1.11,1.38] | 1.11* | [1.02,1.20] |
| **Severity of illness (SOI)[d]** | | | | | | |
| No major or extreme SOI present on admission [Reference] | 1 | (1.00,1.00) | 1 | (1.00,1.00) | 1 | (1.00,1.00) |
| Major SOI | 1.85*** | [1.66,2.05] | 2.98*** | [2.79,3.18] | 2.96*** | [2.80,3.12] |

| | | | | | | |
|---|---|---|---|---|---|---|
| present on admission | | | | | | |
| Extreme SOI present on admission | 3.50*** | [3.13,3.91] | 8.35*** | [7.73,9.02] | 8.32*** | [7.83,8.84] |
| **Hospital strain[g] (% of pre-pandemic average)** | | | | | | |
| 0% to <100% [Reference] | 1 | [1.00,1.00] | 1 | [1.00,1.00] | 1 | [1.00,1.00] |
| 100% to <150% | 1.17*** | [1.10,1.26] | 1.28*** | [1.22,1.34] | 1.29*** | [1.24,1.34] |
| 150% to <200% | 2.67*** | [2.34,3.05] | 2.61*** | [2.41,2.84] | 2.63*** | [2.46,2.81] |
| 200% or higher | 3.39*** | [2.91,3.93] | 3.51*** | [3.19,3.86] | 3.42*** | [3.16,3.69] |
| Principal diagnosis[h] | Yes | | Yes | | Yes | |
| Neighborhood of residence[i] | Yes | | Yes | | Yes | |
| Hospital fixed effect | Yes | | Yes | | Yes | |

Notes:
* p<0.05, ** p<0.01, *** p<0.001
Abbreviations: ICU = intensive care unit; NH = non-Hispanic; SOI = severity of illness; AMC = academic medical center (including affiliates).
[a] < 100 observations were excluded due to missing data.
[b] ICU use is identified based on revenue codes 200, 201, 202, 203,204, 207, 208, 209, 210, 211, 212, 213 and 219 (Weissman et al. 2017).
[c] Non-ICU use is defined as any admissions without revenue codes for ICU use but may include intermediate ICU use.
[d] Severity of illness (SOI) is based on All Patient Refined Diagnosis Related Groups (APR-DRG) classification.
[e] Hospital type is defined as public if a hospital belonged to the NYC H+H system or was state-owned; private (other) hospitals are defined as private hospitals not part of a private AMC system.
[f] Excludes specialty hospitals and hospitals without ICUs (N=6).
[g] Hospital strain is defined as the total number of adult ICU admissions to the hospital in the week prior to and including the patient admission date as a percentage of the hospital's pre-pandemic baseline weekly average ICU admissions.
[h] Principal diagnosis is based on Clinical Classification Software Refined (CCSR) version 2021.1 groupings of ICD-10 diagnostic codes. Top 100 principal diagnoses for hospitalizations resulting in in-hospital death are included. COVID-19 ICD-10 diagnostic code (U.071) went into effect for discharges beginning on April 1, 2020, and does not capture admissions with COVID-19 that resulted in discharges prior to April 1.
[i] Neighborhoods in New York City are defined as Neighborhood Tabulation Areas based on geocoded patient address; separate indicator is included for non-NYC addresses.
Source: New York State Department of Health, Statewide Planning and Research Cooperative System (SPARCS) 2020 inpatient data, 2022 release.

Table S12: Hierarchical logistic regression estimates for inpatient mortality among adults (18+) in NYC, April 1- May 9, 2020[a]

| *Outcome* | **ICU death[b]** (N= 8,713 hosp.) | | **Non-ICU death[c]** (N=59,303 hosp.) | | **Overall hospital death** (N=68,650 hosp.) | |
|---|---|---|---|---|---|---|
| | Odds Ratio | CI 95% | Odds Ratio | CI 95% | Odds Ratio | CI 95% |
| **Patient or admission characteristic** | | | | | | |
| Female | 0.79*** | [0.71,0.88] | 0.94* | [0.88,1.00] | 0.89*** | [0.84,0.93] |
| **Age group** | | | | | | |
| Age 18-39 [Reference] | 1 | [1.00,1.00] | 1 | [1.00,1.00] | 1 | [1.00,1.00] |
| Age 40-64 | 2.21*** | [1.76,2.78] | 3.26*** | [2.53,4.19] | 2.42*** | [2.07,2.83] |
| Age 65 79 | 3.63*** | [2.81,4.70] | 6.82*** | [5.27,8.84] | 4.29*** | [3.63,5.06] |
| Age 80+ | 6.06*** | [4.54,8.09] | 15.33*** | [11.80,19.91] | 8.09*** | [6.83,9.59] |
| **Race/ethnicity** | | | | | | |
| NH White [Reference] | 1 | [1.00,1.00] | 1 | [1.00,1.00] | 1 | [1.00,1.00] |
| Hispanic | 0.9 | [0.76,1.08] | 1.06 | [0.96,1.17] | 1.08 | [0.99,1.17] |
| NH Asian or Pacific Islander | 1.08 | [0.85,1.37] | 1.06 | [0.93,1.21] | 1.12 | [1.00,1.25] |
| NH Black | 0.84 | [0.70,1.02] | 0.87** | [0.78,0.96] | 0.85*** | [0.78,0.93] |
| Other race or ethnicity | 1.18 | [0.98,1.42] | 1.11 | [1.00,1.24] | 1.18*** | [1.08,1.30] |
| **Insurance** | | | | | | |
| Private insurance [Reference] | 1 | [1.00,1.00] | 1 | [1.00,1.00] | 1 | [1.00,1.00] |
| Medicaid | 1.1 | [0.93,1.29] | 1.18* | [1.04,1.34] | 1.09 | [0.99,1.20] |
| Medicare | 1.15 | [0.96,1.38] | 1.41*** | [1.24,1.60] | 1.22*** | [1.11,1.35] |
| Self-pay | 1.13 | [0.68,1.88] | 0.96 | [0.71,1.31] | 0.9 | [0.70,1.16] |
| Other insurance | 1.19 | [0.52,2.71] | 1.91** | [1.21,3.02] | 1.54* | [1.04,2.26] |
| Nursing home admission | 0.77* | [0.62,0.95] | 1.50*** | [1.37,1.65] | 1.23*** | [1.13,1.34] |
| Elixhauser comorbidity score | 1.05*** | [1.04,1.05] | 1.05*** | [1.05,1.06] | 1.06*** | [1.06,1.07] |
| Emergency department | 0.88 | [0.70,1.09] | 0.95 | [0.81,1.13] | 0.89 | [0.78,1.01] |
| **Severity of illness (SOI)[d]** | | | | | | |
| No major or extreme SOI present on admission [Reference] | 1 | (1.00,1.00) | 1 | (1.00,1.00) | 1 | (1.00,1.00) |
| Major SOI | 1.44*** | [1.19,1.73] | 2.31*** | [2.08,2.57] | 2.33*** | [2.13,2.55] |

| | | | | | | |
|---|---|---|---|---|---|---|
| present on admission | | | | | | |
| Extreme SOI present on admission | 2.52*** | [2.08,3.06] | 5.38*** | [4.75,6.09] | 5.58*** | [5.04,6.16] |
| **Hospital type[e, f]** | | | | | | |
| Private - AMC [Reference] | 1 | [1.00,1.00] | 1 | [1.00,1.00] | 1 | [1.00,1.00] |
| Private – other | 1.39* | [1.01,1.92] | 2.56*** | [1.83,3.57] | 2.22*** | [1.66,2.97] |
| Public | 0.98 | [0.71,1.35] | 1.03 | [0.73,1.45] | 1.04 | [0.77,1.40] |
| **Hospital strain (% of pre-pandemic average)[g]** | | | | | | |
| 0% to <100% [Reference] | 1 | [1.00,1.00] | 1 | [1.00,1.00] | 1 | [1.00,1.00] |
| 100% to <150% | 1.60*** | [1.40,1.83] | 1.77*** | [1.63,1.92] | 1.77*** | [1.66,1.90] |
| 150% to <200% | 2.07*** | [1.71,2.52] | 2.55*** | [2.28,2.85] | 2.52*** | [2.29,2.77] |
| 200% or higher | 2.77*** | [2.21,3.47] | 3.64*** | [3.18,4.16] | 3.52*** | [3.14,3.93] |
| cons[hospital] | 1.16*** | [1.06,1.26] | 1.23*** | [1.12,1.36] | 1.18*** | [1.09,1.27] |
| Principal diagnosis[h] | Yes | | Yes | | Yes | |
| Neighborhood of residence[i] | Yes | | Yes | | Yes | |

Notes:
* p<0.05, ** p<0.01, *** p<0.001
Abbreviations: ICU = intensive care unit; NH = non-Hispanic; SOI = severity of illness; AMC = academic medical center (including affiliates).
[a] < 100 observations were excluded due to missing data.
[b] ICU use is identified based on revenue codes 200, 201, 202, 203,204, 207, 208, 209, 210, 211, 212, 213 and 219 (Weissman et al. 2017).
[c] Non-ICU use is defined as any admissions without revenue codes for ICU use but may include intermediate ICU use.
[d] Severity of illness (SOI) is based on All Patient Refined Diagnosis Related Groups (APR-DRG) classification.
[e] Hospital type is defined as public if a hospital belonged to the NYC H+H system or was state-owned; private (other) hospitals are defined as private hospitals not part of a private AMC system.
[f] Excludes specialty hospitals and hospitals without ICUs (N=6).
[g] Hospital strain is defined as the total number of adult ICU admissions to the hospital in the week prior to and including the patient admission date as a percentage of the hospital's pre-pandemic baseline weekly average ICU admissions.
[h] Principal diagnosis is based on Clinical Classification Software Refined (CCSR) version 2021.1 groupings of ICD-10 diagnostic codes. Top 100 principal diagnoses for hospitalizations resulting in in-hospital death are included. COVID-19 ICD-10 diagnostic code (U.071) went into effect for discharges beginning on April 1, 2020, and does not capture admissions with COVID-19 that resulted in discharges prior to April 1.
[i] Neighborhoods in New York City are defined as Neighborhood Tabulation Areas based on geocoded patient address; separate indicator is included for non-NYC addresses.
Source: New York State Department of Health, Statewide Planning and Research Cooperative System (SPARCS) 2020 inpatient data, 2022 release.
